# Assessing early detection ability through spatial arrangements in environmental surveillance for poliovirus: a simulation-based study

**DOI:** 10.1101/2024.08.17.24312151

**Authors:** Toshiaki R. Asakura, Kathleen M. O’Reilly

## Abstract

Detecting the circulation of poliovirus in its early stages is paramount for swift public health action. Environmental surveillance (ES) is the testing of sewage samples for the presence of poliovirus and has the potential to enhance early detection capabilities. However, the establishment and maintenance of ES sites incur costs and necessitate human resources, underscoring the consideration of ES scale and site selection. Here, we aim to assess the early detection ability of ES by varying the number and location of ES sites using the simulation-based approach. We developed a stochastic meta-population model among unimmunised children aged under 5 years old and used geographic and demographic characteristics of South Africa as a case study of a non-endemic country, assuming a single introduction of wild poliovirus serotype 1. We constructed six scenarios by combining three importation risk distributions (predicated on population size, approximations of international inbound travel volume and border crossing volume) with two ES site layout strategies: one proportionate to population size and another focusing on importation risk via land border crossings. We showed that a modest number of strategically positioned ES sites can achieve a high early detection ability when assumed importation risks were geographically confined. In contrast, when importation risks were dispersed, the effectiveness of ES was diminished. Our sensitivity analyses suggested that the strategy to implement ES across large areas with low sampling frequency consistently resulted in a better early detection ability against various importation scenarios than one to implement ES in limited areas with high sampling frequency. Although we acknowledge the challenges of translating our simulated outcomes for real-world situations, our study has implications for deciding the scale and site selection of ES.

**Author summary:** Poliovirus causes paralytic poliomyelitis, a terrible and debilitating disease. Only a small number of infections with poliovirus result in disease. Early detection of these infections can inform public health decision-makers to trigger interventions such as vaccination campaigns. Environmental surveillance (ES) is able to detect poliovirus circulation by capturing virus shedding (even from asymptomatic people) in wastewater. Strategic geographical placements of ES sites may enhance early detection capabilities, but there has been limited exploration of factors that might affect early detection. Here, we investigated the relationship between the scale and site selection of ES and early detection ability by simulating the introduction, spread and detection of poliovirus among unimmunised children aged under 5 years old. We used geographic and demographic characteristics of South Africa as a case study of a non-endemic country, assuming an introduction of wild poliovirus serotype 1. We found that the informed ES placement achieved a high early detection ability when importation risks were geographically limited. In contrast, when importation risks were dispersed, the ES effectiveness was diminished. Our sensitivity results suggested the strategy to expand ES sites was robust against various importation scenarios and better than the strategy to merely increase sampling frequency in limited locations.

## Introduction

Global concerted efforts toward polio eradication have achieved a drastic reduction in the number of poliomyelitis cases [1] and cooperated surveillance systems contributed to this achievement [2]. Patients with paralytic poliomyelitis are detected through syndromic surveillance, referred to as acute flaccid paralysis (AFP) surveillance, but a tiny portion of infections can be detected due to a very low paralysis-to-infection rate. It has been estimated that for every 200 wild poliovirus serotype 1 (WPV1) infections there will be one paralytic case [3]. To improve surveillance to rule out local transmission of poliovirus, sewage sampling, which is referred to as environmental surveillance (ES), has been developed. After the withdrawal of the trivalent oral polio vaccine (OPV) in 2016 and the emergence of vaccine-derived poliovirus serotype 2 (VDPV2) outbreaks [4,5], ES has been a vital complementary surveillance tool for polio eradication [6–8], especially for the detection of cryptic circulation in subnational areas of endemic countries, and detection of importation or confirmation of polio-free status in non-endemic countries.

Detection of poliovirus circulation through ES can trigger swift public health actions to contain outbreaks [9]. Recent examples include the detection of VDPV2 circulation first through the ES in the US [10,11] and the UK [12], which enabled public health authorities to conduct active case finding, supplementary immunisation activity and social mobilisations. On the other hand, delays in detection have been linked to a large number of cases during outbreaks [13]. The long reporting delays of AFP surveillance have been attributable to a delay in sample collection, transport, culture and sequencing as well as the time required to ship collected samples to other countries due to the lack of facilities in resource-limited settings [13,14].

Expanding ES sites can enhance early detection capabilities, but the establishment and maintenance of ES sites incur costs and necessitate human resources [15]. To operate ES effectively, the quality assessment is essential, which comprises the appropriateness of sampling site locations [16], importation risk assessment [17], ES-covered population size, non-enterovirus detection [18], and the quality of sample handling and sample processing. Although guidelines for the implementation of ES have been developed by the Global Polio Eradication Initiative (GPEI) [19], specific guidance on the number and location of ES sites is still lacking due to uncertainties in available resources.

Quantitative evidence of the early detection ability of ES is needed to design ES layout strategies at the national or subnational level. One study empirically investigated the early detection capabilities using poliovirus genome data in Pakistan, showing that ES can detect the circulation of specific genotypic clusters before AFP surveillance in nearly 60% of sampled clusters [20]. From the perspective of a mathematical modelling approach, one seminal paper quantified the simulated cumulative probability of detecting poliovirus circulation through each AFP surveillance and ES [21], and one paper broadly examined the lead time of the first detection through ES over other surveillance systems assuming various pathogen characteristics [22]. Another study theoretically investigated optimal sampling frequency against emerging pathogens, considering a balance between sampling costs and disease burden [23].

There remains a gap in understanding the quantitative relationship between the ES early detection ability and spatial arrangements of ES sites. While laboratory facilities may be constrained by capacity, there is flexibility in the selection of ES sites and we aim to address this point using a simulational approach. Here, we utilised geographic and demographic characteristics of South Africa (where a polio-free status has been maintained since 1989 [24]) as a case study of a non-endemic country. We employed the stochastic meta-population framework (following the basic model structure proposed by Ranta et al. [21]) among unimmunised children aged under 5 years old to assess the first detection timing through AFP surveillance and ES. We assumed that a single patient with WPV1 was imported to a specific ‘patch’ (a unit of a spatial grid in the meta-population framework) according to the importation risk distribution. Our study setting was motivated by the WPV1 importation event of 2022 into Mozambique, a neighbouring country to South Africa, implying a non-negligible risk of WPV1 importation into South Africa [25].

By conducting simulations, we aim to explore the quantitative relationship between the number and location of ES sites and the early detection capabilities of ES over AFP surveillance.

## Results

### Study setting

The overview of our study setting is summarised in Fig 1 and a schematic representation of our model is shown (Fig S3 in S1 text). We refer to a patch as a unit square at 20km spatial resolution (i.e. 400 km^2^), matched with the actual geographical location within the meta-population framework. The ES site was assumed to be placed within a single patch, covering 25% of the population of that patch. We chose 25% for a patch-level ES population coverage by considering the observed national ES population coverage and district-level ES population coverage (Table S4-5 and Fig S8 in S1 text). Our SEIR model is comprised of three components: transmission model, AFP surveillance model and ES surveillance model (Fig S3 in S1 text). We assumed the probability of detecting poliovirus circulation through ES was proportional to the incidence rate of infected individuals. We define ‘LT’ as the lead time of the first detection (in days) through the ES over AFP surveillance and seven detection patterns were considered: ‘No detection’, ‘AFP only detection’, earlier detection through AFP surveillance than ES (i.e. ‘<-60 LT’, ‘-60 ∼ −1 LT’), earlier detection through ES than AFP surveillance (i.e. ‘0 ∼ 59 LT’, ‘≥60 LT’) and ‘ES only detection’ (Fig 1B&C). We used the proportion of each detection pattern among simulations with poliovirus detections (i.e. excluding the ‘no detection’ pattern) as the main output of simulation results.

**Fig 1.**
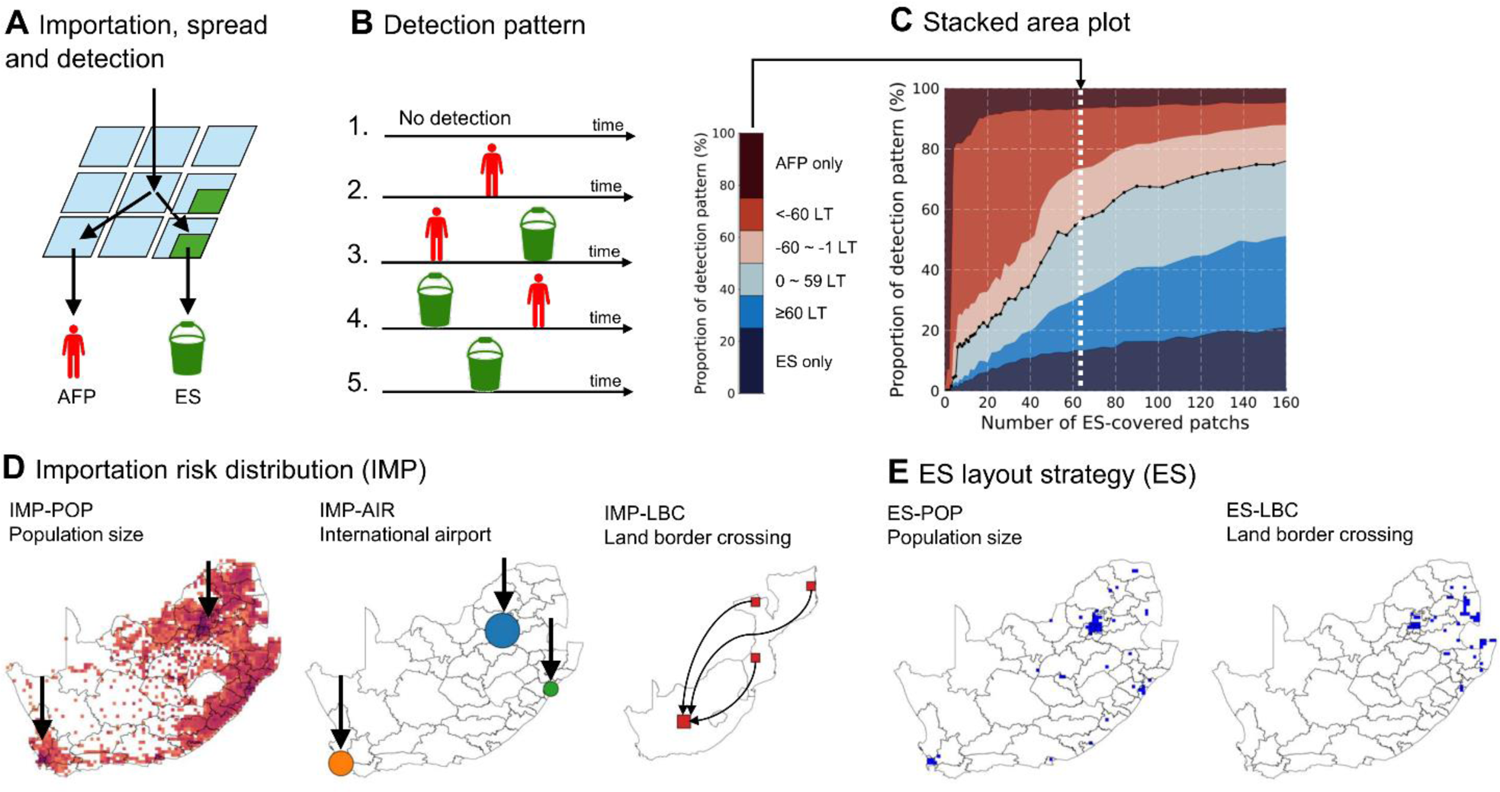
Schematic illustration of study settings. **(A)** Simulated importation of wild poliovirus serotype 1 to South Africa, spread described by the meta-population model, and detection through acute flaccid surveillance (AFP) and environmental surveillance (ES). AFP surveillance is carried out across all patches whereas ES operations are confined to selected patches (in green). A patch-level ES population coverage is assumed to be 25%. **(B)** Five detection patterns: i) No detection, ii) AFP only detection, iii) earlier detection through AFP surveillance than ES, iv) earlier detection through ES than AFP surveillance v) ES only detection. Detection patterns of iii) and iv) are further classified based on the lead time (LT) of the first detection through ES over AFP surveillance. **(C)** Stacked area plot of the proportion of each detection pattern against the number of ES-covered patches. **(D)** Three importation risk distributions: i) IMP-POP corresponds to the ‘Population size’-based importation risk distribution and assumes importation risk is proportional to the population size of each patch (indicated by orange-red colouration); ii) IMP-AIR corresponds to the ‘International airport’-based importation risk distribution and assumes an importation risk is shaped by international inbound travel volume in 2019 and approximated movement from airports. Each circle denotes the international airport with a size proportional to the international inbound travel volume: Blue, O.R. Tambo International Airport; Orange, Cape Town International Airport; Green, King Shaka International Airport; iii) IMP-LBC corresponds to ‘Land border crossing’-based importation risk distribution and assumes importation risk is proportional to human mobilisation from outbreak countries, in this study Mozambique. **(E)** Two ES site layout strategies: i) ES-POP denotes ‘Population size’ where allocation of sites is ordered by population size in each patch; ii) ES-LBC denotes ‘Land border crossing importation risk’ where ES sites are placed according to assumed international travel volume via land border crossings.

Preliminary analysis illustrated that the first detection timing through ES was largely influenced by the assumed route of poliovirus introduction, and ES site layout. Therefore we prepared three importation risk distributions and two ES site layout strategies, totalling 6 scenarios when combined (Fig 1D&E, S7 in S1 text and S1-2 Video). For example, IMP-POP denotes the population size-based importation risk distribution and assumes that an importation risk is proportional to the population size of each patch (Fig 1D and S7A in S1 text). The ES site layout strategy determines the sequence in which patches are covered by the ES. For example, ES-LBC denotes land border crossing importation risk-based ES site layout strategy and assumes that we first cover patches with a high importation risk via land border crossing from Mozambique (Fig 1E, S8C in S1 text and S2 Video). Six scenarios are expressed with the combinations of importation risk distribution and ES layout strategy as the following: IMP-POP/ES-POP, IMP-AIR/ES-POP, IMP-LBC/ES-POP, IMP-POP/ES-LBC, IMP-AIR/ES-LBC, and IMP-LBC/ES-LBC.

### A modest number of targeted ES site implementations can achieve high simulated early detection probability

The proportion of each detection pattern was visualised as the stacked area plot against the number of ES-covered patches for 6 scenarios (Fig 2). We truncated the number of ES-covered patches at 160 sites even though the total number of patches for all of South Africa was 1502. The full scale of the figure can be found in Fig S12 in S1 text, in which the x-axis represents the national ES population coverage. When importation risks were concentrated in confined patches (e.g. IMP-AIR and IMP-LBC) and an ES site layout strategy was prioritised to cover those high importation risk areas (e.g. IMP-AIR/ES-POP and IMP-LBC/ES-LBC, shown in Fig 2B and 2F), 6–8 ES-covered patches were sufficient to achieve 50% simulated early detection probability. In comparison, in the scenario where the ES site layout strategy failed to prioritise patches with high importation risks (e.g. IMP-LBC/ES-POP and IMP-AIR/ES-LBC, shown in Fig 2C and 2E), covering only 10 to 20 patches by ES resulted in low simulated early detection probability.

**Fig 2.**
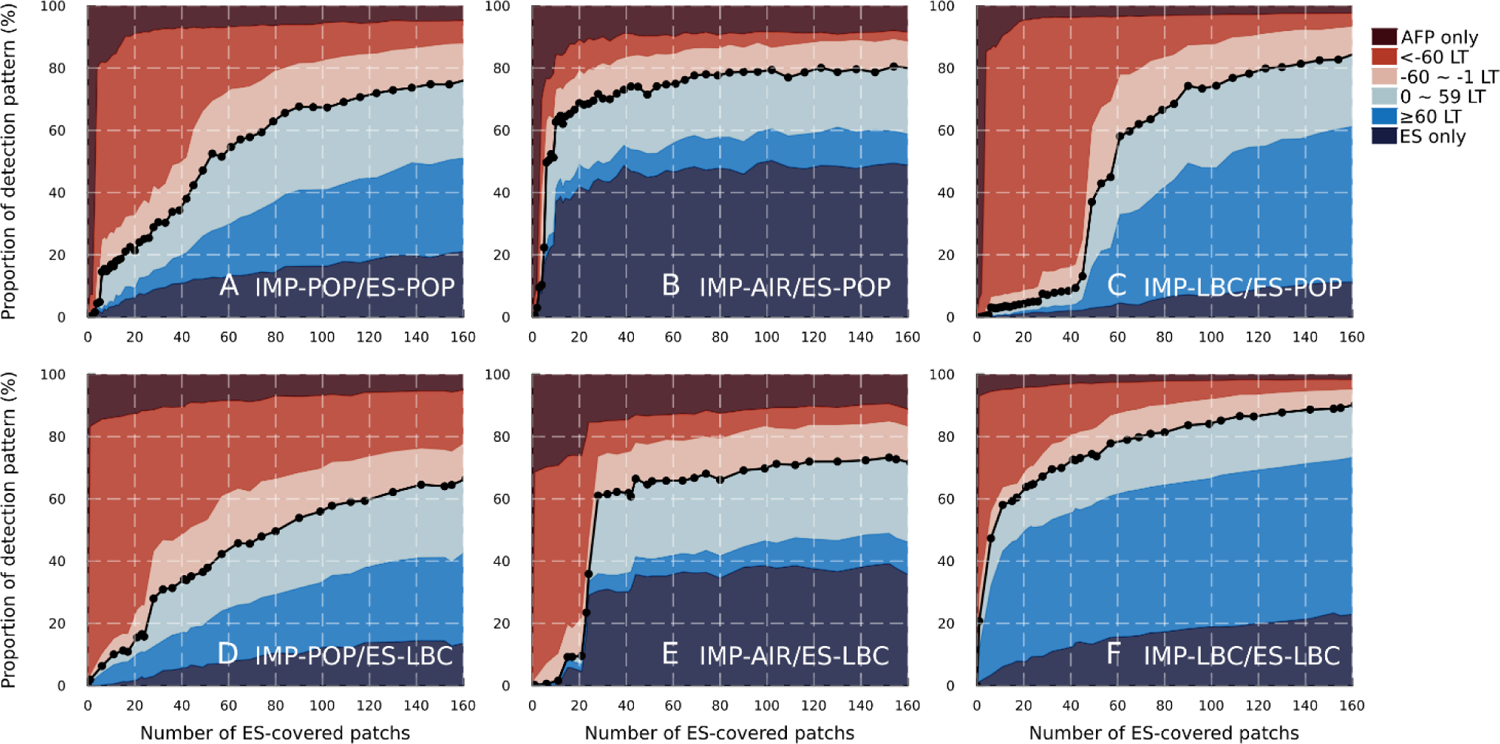
Proportion of each detection pattern (%) against the number of ES-covered patches for 6 scenarios. The blue-coloured area under the black dotted lines represents the simulated early detection probability, consisting of the early detection through ES over AFP surveillance and the ES only detection pattern. It is noted that the maximum number of ES-covered patches is 1502 and the x-axis is truncated at 160. IMP-POP, ‘Population size’-based importation risk distribution; IMP-AIR, ‘International airport’-based importation risk distribution; IMP-LBC, ‘Land border crossing’-based importation risk distribution; ES-POP, ‘Population size’-based ES site layout strategy; ES-LBC, ‘Land border crossing importation risk’-based ES site layout strategy. LT denotes the lead time of the first poliovirus detection through ES over AFP surveillance.

According to the wastewater plant data provided by the National Institute of Communicable Disease in South Africa on 27 November 2023, ES for poliovirus was operating at 17 wastewater plants, covering ∼11.3% of the national population (if the wastewater-served population was imputed with the median value for two wastewater plants; see S1 text for details). Under our model assumptions, the closest number of ES-covered patches to match the current ES capacity in South Africa (i.e. the number of ES-covered patches required to achieve 11.3% national ES population coverage) was 58 for ES-POP and 154 for ES-LBC. To investigate the impact of the exclusion of the ‘no detection pattern’ on the proportion of detection patterns, we visualised those proportions including the ‘no detection’ pattern in Fig S13 in S1 text. We found significant differences in the simulated probability of at least one detection of poliovirus either through ES or AFP surveillance (coloured areas), which would be attributable to heterogeneity in the effective immunisation proportions.

### Sensitivity analysis showed sampling frequency and ES sensitivity were key parameters for enhancing early detection ability

We conducted a sensitivity analysis of the basic reproduction number (*R*_0_), travelling rate between patches (α), sampling frequency, ES sensitivity, and patch-level ES population coverage (*p*_c_) for the IMP-POP/ES-POP scenario (Fig 3 and 4). The basic reproduction number of poliovirus (*R*_0_) did not influence the simulated early detection probability in the entire region of the number of ES-covered patches, whereas a lower basic reproduction number resulted in a large proportion of AFP only or ES only detection patterns. This tendency was consistent with the sensitivity analysis under the single patch setting (Fig S10 in S1 text).

**Fig 3.**
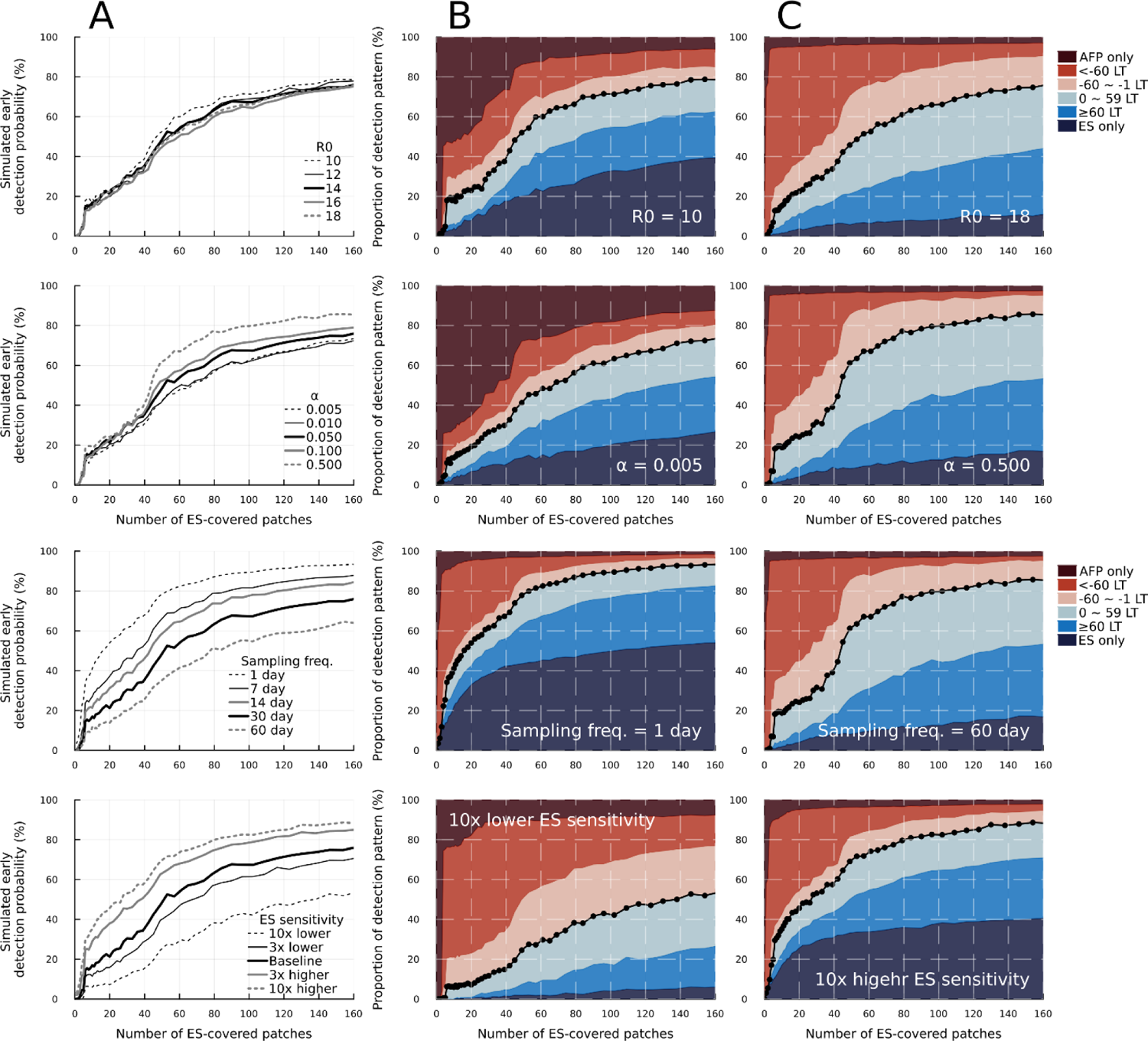
Sensitivity analysis of the basic reproduction number (*R_0_*), travelling rate between patches (*α*), sampling frequency and ES sensitivity for the IMP-POP/ES-POP scenario. (A) Simulated early detection probability for different parameters against the number of ES-covered patches. Thick black lines represent results with the same parameter value as in the main analysis. (B, C) Stacked area plot for the smallest and largest sensitivity parameter values. LT corresponds to the lead time of the first poliovirus detection through ES over AFP surveillance and ‘sampling freq.’ corresponds to the sampling frequency.

**Fig 4.**
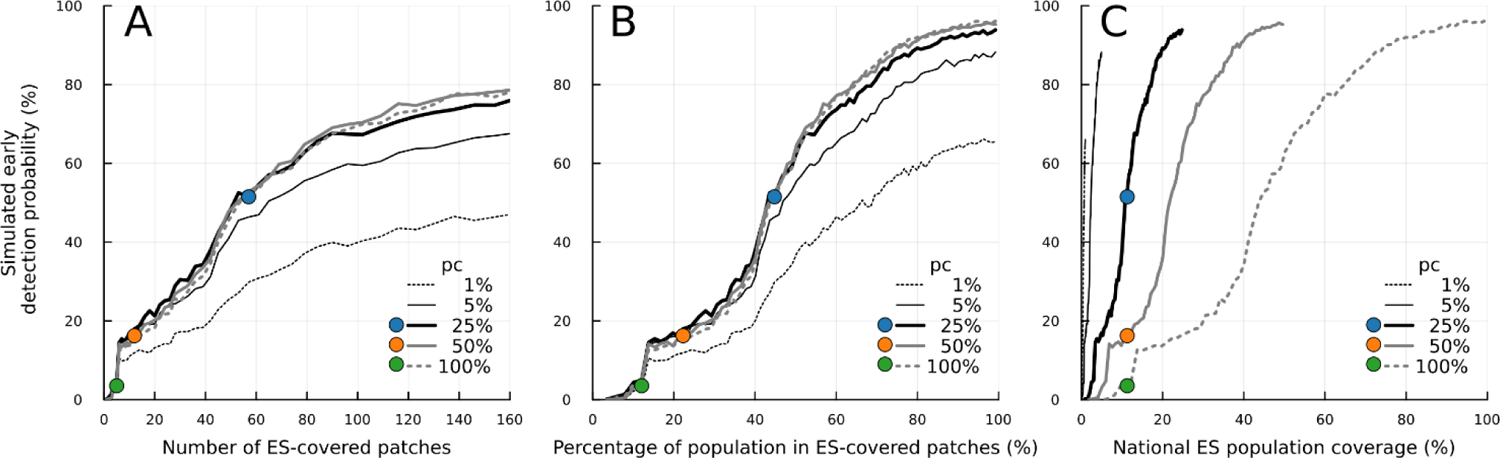
Sensitivity analysis of the patch-level ES population coverage (*p_c_*) for IMP-POP/ES-POP scenario. (A, B, C) Simulated early detection probability is plotted (A) against the number of ES-covered patches, (B) against the percentage of the population in ES-covered patches, and (C) the national ES population coverage. Black lines represent simulation results for each of *p_c_* and the number of ES-covered patches. The coloured data points in (A-C) represent simulations where the simulated national ES population coverage aligned with the observed national ES population coverage in South Africa (11.3%), under *p_c_* of 25% (blue), 50% (orange) and 100% (green). The national ES population coverage is given by the product of *p*_c_ and the percentage of the population in ES-coverage patches. It is noted that the maximum number of ES-covered patches is 1502 and the x-axis for (A) is limited to a maximum value of 160.

The simulated early detection probability remained consistent across different travelling rates (*α*) for a small number of ES-covered patches (i.e. up to 40 patches) whereas increasing the value of *α* by tenfold, from the baseline value of 0.050 to 0.500, yielded a 15% increase in simulated early detection probability after the number of ES-covered patches exceeded 50. Notable disparities were consistently observed when focusing on the proportion of AFP only or ES only detection patterns. Setting high travelling rates resulted in smaller proportions of ES only detection patterns whereas setting low travelling rates resulted in higher proportions. Both sampling frequency and ES sensitivity (which is governed by two parameters of lognormal distribution) influenced the simulated early detection probability in the order of 10 to 25%, and these differences were consistently observed across the entire region of the number of ES-covered patches. It is noted that either with the higher sampling frequency or higher ES sensitivity, ES only detection pattern accounted for more than 30% among simulations excluding no detection pattern.

### Simulated early detection ability largely depends on the choice of patch-level ES population coverage (*p*c)

We explored the impact of the patch-level ES population coverage (*p*_c_) on simulated early detection ability for the IMP-POP/ES-POP scenario. The parameter *p*_c_ modulated the balance between the concentration of ES placement in a single patch and the dispersion of ES sites across patches under a fixed national ES population coverage. The simulated early detection ability remained nearly consistent for *p*_c_ values greater than 5%, given the same number of ES-covered patches (Fig 4A). Considering the observed district-level ES population coverages in South Africa all exceeding 10% (with a median of 22.5%), the early detection ability would be robust despite large variations in ES coverage in those districts (Table S3 in S1 text).

Since the higher *p*_c_ value leads to a larger ES-covered population with limited ES-covered sites, we plotted the coloured points representing the simulation results where the simulated national ES population coverage matched the observed coverage (Fig 4A). Under such a constraint, when *p*_c_ =100%, only 5 sites were required to achieve the observed national ES population coverage but the combined sensitivity of these sites resulted in a low simulated probability of early detection (Fig 4A, green circle). Lower *p*_c_ values correspond with many ES-covered patches, which in turn increased simulated early detection probability (Fig 4A, blue circle).

To further illustrate the role of the patch-level ES population coverage parameter, the same simulation results are displayed in two different x-axes. First, we employed the percentage of the population in ES-covered patches, which was calculated by the sum of the population in ES-covered patches divided by the total population size (Fig 4B). It is noted that (1-*p*_c_)% of the population was not covered by ES but counted in the denominator. The tendency of simulated early detection probability was similar to Fig 4A. Second, we considered the national ES population coverage as the x-axis, which was calculated by multiplying *p*_c_ with the “percentage of the population in ES-covered patches” to correctly account for the population covered by ES. Even though the same national ES population coverage was maintained (Fig 4C, coloured points), the simulated early detection ability largely depended on the choice of *p*_c_. This large variation implies the national ES population coverage would be an unreliable measurement to evaluate the spatial arrangements of the ES sites for the early detection ability. This tendency was consistent across the sensitivity analyses under the other 5 scenarios (Fig S14-15 in S1 text).

### Correlations between simulated early detection probability and weighted average minimum distance to ES-covered patches

We explored more parsimonious assessment measurements for the ES site layout across the countries since our stochastic simulations required a long computation time. We calculated the average minimum distance from patches with importation to ES-covered patches weighted by outbreak probability. The outbreak probability in the present study is defined as the probability of 10 or more infections occurring given a single introduction, considering the effective immunisation proportion (EIP) in each patch (Fig S9 in S1 text). This metric can be calculated if importation risks and ES site layout are available. Intuitively, early detection probability is likely to be higher if ES-covered patches are placed near locations for importation. In this exercise, we wanted to explore whether assumptions on importation and ES placement consistently influenced this property.

The relationship between simulated early detection probability and weighted average minimum distance showed a shape similar to an exponential curve for the IMP-POP and IMP-LBC scenarios (Fig 5A&B). Once the simulated early detection probability surpassed 50%, this relationship transitioned towards a nearly linear trend. Although the shape for these relationships was similar for ES-POP and ES-LBC scenarios, the scale of weighted average minimum distance was different.

**Fig 5.**
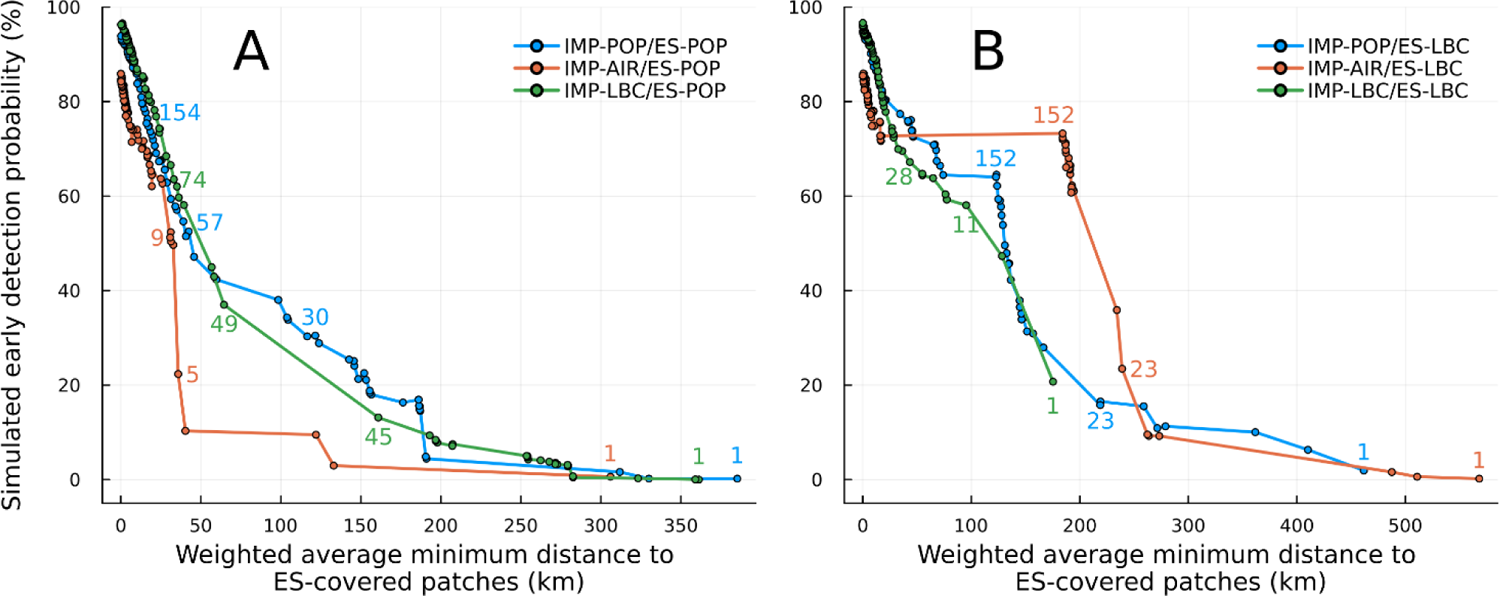
Relationship between the weighted average minimum distance to ES-covered patches and simulated early detection probability (%). (A) Under ES-POP scenarios. (B) Under ES-LBC scenarios. Minimum distances to ES-covered patches were weighted by importation risk and outbreak probability of 10 or more infections occurring, considering the effective immunisation proportion. The coloured numbers next to points correspond to the number of ES-covered patches for each importation risk distribution.

In contrast, the relationship between simulated early detection ability and weighted average minimum distance showed an irregular pattern in IMP-AIR scenarios. We observed a sharp increase in simulated early detection probability despite a small difference in weighted average minimum distance in both ES-POP and ES-LBC. Conversely, only a small difference in simulated early detection probability was present despite a large difference in weighted average minimum distance. Notably, in the IMP-AIR/ES-LBC scenario, when the number of ES-covered patches exceeded 152, there was no increase in simulated early detection probability despite a more than 100km decline in the weighted average minimum distance to ES-covered patches.

## Discussion

In this study, we assessed the quantitative relationship between early detection ability and the scale and locations of ES sites by employing a meta-population framework. We simulated the importation, spread and detection process, varying the number of ES-covered sites under different importation risk distributions and ES site layout strategies. By applying stochastic simulations, we successfully considered the partial detection patterns (i.e. ES only detection or AFP surveillance only detection) to align with real-world observations. The results provided here illustrated the potential of strategic positioning of ES sites to enhance early detection capabilities and clarified key ES-related parameters to be considered. Our results also highlight the importance of poliovirus importation risk assessment and how infectious disease surveillance should be tailored to perceived threats.

We found that simulated early detection probability exhibited a monotonic increase with the number of ES sites, but distinct variations in slope and plateau points were observed across six different scenarios. When importation risks were concentrated in confined patches, the modest number of strategic and targeted ES positioning can be highly efficient in the early detection of poliovirus circulation. Conversely, if importation risk was geographically dispersed, the effectiveness of ES was diminished, and many ES sites were required for a high early detection ability. It is noted that in our simulation study, we did not consider delays in reporting from the onset of AFP or delays in processing environmental samples. The reporting delays in patients with AFP were substantial in resource-limited settings, which could be around 29–74 days [22], and those delays should be considered in practice.

Our sensitivity analysis showed large variations in simulated early detection probability depending on the patch-level ES population coverage given the same national ES population coverage, which poses the challenge of translating our simulation results into the practical ES implementation strategy. Although national ES population coverage can be easily calculated, this measurement would poorly reflect early detection ability for the ES layout unless a patch-level ES population coverage is precisely specified. On the other hand, it is difficult to relate a patch in our simulations to real-world geographical settings. We roughly assumed the spatial resolution of each patch was equal to 20 x 20 km (i.e. 400 km^2^) regardless of urban and rural areas and made homogenous mixing assumptions within a patch. This assumption led to unreasonable model-based conclusions that ES sites should cover as small a proportion of the population within a patch as possible and as many patches as possible, resulting in high early detection probability with low national ES population coverage (requiring fewer resources) (Fig 4C). This result is an artefact of assuming that all individuals within a patch mix homogeneously, and therefore a tiny ES area coverage within a patch would have an unreasonably high probability of detecting poliovirus. Thus, crucial information for connecting models with real-world settings is needed to identify areas where homogenous mixing is held, and in other words, to identify the fragmentation level of patches, as is historically pointed out by several authors [26–29]. One study of COVID-19 provided some insights into the homogenous mixing assumptions. This study used virus genome data with corresponding resident addresses in Danne, Scotland, suggesting around a 5km radius circle was well mixed in terms of genetic distance [30]. An alternative approach would have been to reduce patch size, for example to 5 x 5 km, which may align better with the size of ES catchments and assumptions on homogenous mixing. However, this procedure results in a large increase in computation time, and the chatchment sie of environmental sites is typically unavailable in low- and middle-income settings and we are therefore trying to align simulations with a uncertain value. An improved understanding of catchment sizes in real-world settings is warranted because our study illustrates the sensitivity to assumptions on catchment (or patch) size on early detection ability.

The sampling frequency is a parameter of interest to optimise the ES site layout [17,23]. By employing a daily sampling strategy in our study, the ES early detection ability increased significantly. However, this strategy resulted in an increased proportion of the ES only detection pattern, which might cause overacting against such an importation that does not lead to secondary infections. Assuming an equal number of environmental samples can be taken for each month, extending ES sites can be more efficient in improving early detection ability and be robust against various importation risk distributions. For example, if we assumed 20 ES-covered sites and monthly sampling for the IMP-POP/ES-POP scenario, doubling sampling frequency led to a 10% increase in simulated early detection probability while doubling ES-covered sites resulted in the same increase and more resilience against multiple importation risk distributions. Moreover, the latter strategy can minimise the proportion of the ES only detection pattern. One concerning point is that expanding ES sites requires much more cost and additional human resources for transportation than increasing sampling frequency [16,31].

The basic reproduction number (*R*_0_) for poliovirus is difficult to determine due to the small number of reported paralytic cases per outbreak and the impact of changes in hygiene. Our sensitivity analysis of *R*_0_ showed small differences in simulated early detection probabilities, supporting the high robustness of results in the present study. Our assumed *R*_0_ for the main analysis was based on a review of transmissibility by Fine et al. from 1999 [32], and the hygiene level in the African continent has since greatly improved. We therefore expect the current *R*_0_ value in South Africa will be lower, but the sensitivity analysis would be well within the bounds of potential *R*_0_ values. One study quantitatively investigated the early detection ability varying *R*_0_ and other pathogen characteristics using a branching process [22] and found that the lead time for wastewater surveillance was different depending on *R*_0_. Two reasons can be considered. First, our model assumed an effective reproduction number of around one by considering vaccination coverage. So, the variation of the effective reproduction number was smaller compared to the range of *R*_0_ investigated in Andrew, et al. [22]. Second, we considered ES only detection patterns as early detection, which did not happen in the branching process model.

Our study is not free from limitations. First, we considered the country where OPV and inactivated polio vaccine (IPV) are routinely administered. The large outbreak of VDPV2 was caused by switching from trivalent OPV to bivalent OPV [33], and many countries are planning to cease OPV usage except for outbreak response [34,35]. Moreover, in many developed countries, IPV is only included in their routine immunisation schedule. Those vaccinated with only IPV could spread poliovirus to others due to lack of mucosal immunity and could be detected through ES, but they would be less likely to develop AFP due to the humoral immunity, implying increased utility of the ES compared to our study (where an IPV-OPV schedule is assumed). Second, we only focused on WPV1 and did not consider other serotypes (such as cVDPVs) or transmission from immune-compromised individuals shedding (iVDPVs).

Third, we limited transmission dynamics in children under 5 years old, assuming those aged 5 or more were completely immunised. However, the reported age distribution of poliomyelitis patients is skewed towards older groups in non-endemic countries [36], and multiple reasons for being unvaccinated were considered such as migration, poverty and conflict [37–39]. We ignored those pocket populations considering the size of that population and the paucity of historical vaccine coverage data in South Africa. Fourth, our importation risk distributions were essentially based on the population size of each patch and the approximated mobilisation pattern by the radiation model, which could make the ES performance better when compared to reality. Quantitative data about international traveller movement from airports and border crossing populations could improve predictions that support risk assessment. Furthermore, research on the importation pathway is demanding for prevention, detection and response. The importation routes to reported outbreak sites were often unknown and one to three years of cryptic circulation was suspected for some outbreaks [25]. Lastly, parameters for ES sensitivity, which was described by the lognormal distribution, were estimated using the wastewater surveillance data for COVID-19 in the US [40].

Considering differences in virus shedding characteristics between poliovirus [41] and COVID-19 [42,43] as well as differences in ES site system and quality between the US and other low- and middle-income countries, the ES sensitivity will be inherently different, underscoring the need for region-specific data to assess site-specific ES sensitivity.

In conclusion, several countries are planning to initiate, expand and optimise the ES for poliovirus. We varied the number and location of the ES sites under different importation risk distributions to quantify the ES early detection ability over AFP surveillance. Our results showed that risk-targeted ES site layout could achieve high early detection capabilities. Further research is required to optimise resources for the ES to monitor the progress toward polio eradication.

## Material and Methods

### Data

We collated population data for children under 5 years old and for all age groups in 100m spatial resolution in South Africa and Mozambique from WorldPop, respectively [44]. We aggregated each dataset to obtain a 20km spatial resolution and called each unit of area a ‘patch’ within a meta-population framework. We removed patches with less than 100 children under 5 years old for computational efficiency, resulting in 1502 patches in South Africa (Fig S1 in S1 text).

We collated the district-level vaccination coverage from the Expanded Programme on Immunisation National Coverage Survey Report 2020 in South Africa [45]. South Africa included both bivalent OPV and IPV in their routine immunisation schedule. In South Africa’s vaccine schedule, OPV is administered at birth and 6 weeks after birth, and IPV is administered as a part of the hexavalent vaccine (HEXA) at 6 weeks, 10 weeks, 14 weeks, and 18 months after birth.

To quantitatively validate the choice of patch-level ES population coverage (*p*_c_), we obtained the wastewater plant information about the location and their served-population size from the National Institute for Communicable Diseases (NICD). The details of the wastewater data description and validation process can be found in Table S3-4 and Fig S8 in S1 text.

### Proportion of immunised children under 5 years old

We assumed those vaccinated with at least one IPV also completed two doses of OPV in our simulation, and classified children into immunised and unimmunised ones. Immunised children are assumed to have no susceptibility and transmissibility because of mucosal immunity induced by OPV and they are assumed not to develop AFP because of serum antibody induced by IPV. Therefore, we removed immunised children from the transmission dynamics. Since the last known polio patient in South Africa was documented in 1989 [24], immunity from natural infection was not considered.

We calculated the proportion of children under 5 years old who were effectively immunised, which we called the effective immunisation proportion (EIP), with the district-level coverage of IPV and assumptions of vaccine effectiveness against transmission. Let *VE*_p_ be the vaccine effectiveness per dose (0.63 against serotype 1 [46–48]) and the probability of immunisation after *n* doses of IPV (*VE*_n_) is calculated as *1 – (1-VE*_p_)^n^. Considering the IPV coverage for *n*th dose (*C*_v,n_), the EIP can be estimated considering *VE*_n_ and the IPV coverage for *n*th dose (*C*_v,n_), which is described by *C*_v,4_*VE*_4_ + (*C*_v,3_ – *C*_v,4_)*VE*_3_ + (*C*_v,2_ – *C*_v,3_)*VE*_2_ + (*C*_v,1_ – *C*_v,2_)*VE*_1_. When *C*_v,n_ < *C*_v,n+1_, we assumed *C*_v,n_ - *C*_v,n++1_ to be zero. We calculated the district-level EIPs (Table S1 and Fig S2 in S1 text) and assigned them as the corresponding patch-level EIP. The average EIP weighted by the population size of children under 5 years old in each patch in South Africa was 91.9%.

### Modelling framework

We constructed a stochastic meta-population model among unimmunised children under 5 years old considering the detection process of poliovirus through AFP surveillance and ES. The model comprises three components: the transmission model, the AFP surveillance model, and the ES model. In each simulation, we randomly chose one patch to introduce WPV1 proportional to the assumed importation risks of each patch and ran the stochastic SEIR meta-population model until one of the following criteria was met: the first poliovirus was detected through both surveillance, no polio patients were present, or 3 years had passed from the beginning of the simulation.

Since the first detection timing through ES is largely influenced by the distribution of importation risks and ES site layout strategy, we prepared three importation risk distributions and two ES site layout strategies as follows (Fig 1, Fig S7-8 in S1 text and S1-2 Video):

– IMP-POP denotes ‘Population size’-based importation risk distribution and assumes the importation risk of each patch is proportional to the population size of each patch.
– IMP-AIR denotes ‘International airport’-based importation risk distribution and assumes the importation risk of each patch is proportional to international inbound travel volume in 2019 and further considers mobilisation from airports.
– IMP-LBC denotes ‘Land border crossing’-based importation risk distribution and assumes the importation risk of each patch is proportional to travelling volume from Mozambique, which is approximated by the radiation model.
– ES-POP corresponds to ‘Population size’-based ES site layout strategy, and assumes the ES is implemented in the descending order of population size of each patch.
– ES-LBC corresponds to ‘Land border crossing importation risk’-based ES site layout strategy and assumes the ES is first implemented in a patch with a high importation risk via land border crossing from Mozambique.

We considered the combination of each importation risk distribution and ES site layout strategy, totalling 6 scenarios: IMP-POP/ES-POP, IMP-AIR/ES-POP, IMP-LBC/ES-POP, IMP-POP/ES-LBC, IMP-AIR/ES-LBC, and IMP-LBC/ES-LBC. We included the land border crossing scenarios to assess the early detection ability against the poliovirus introduction in rural settings with an informed ES layout strategy (i.e. IMP-LBC/ES-LBC scenario).

In each scenario, we ran 10,000 simulations and calculated the proportion of each detection pattern, varying the number of ES-covered patches. All analyses were performed in Julia v1.8.3.

### Transmission model

We employed a stochastic meta-population SEIR model among unimmunised children under 5 years old (Fig S3 in S1 text). The SEIR model simulates the process in which susceptible individuals (S) get infected, passing through a latent period (E) and infectious period (I), and then recover from infection (R). The daily hazard rate for newly infected individuals in patch *i* at day *t* is expressed as

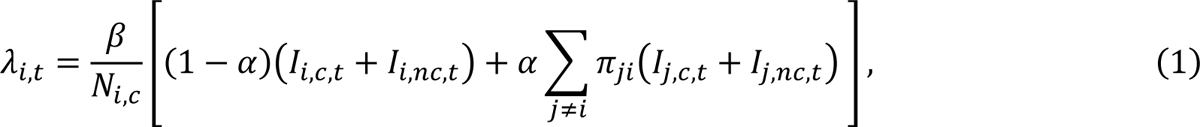

where *β* corresponds to the transmission rate, *N*_i,c_ is the population size of children under 5 years old of patch *i* regardless of immunity status, *α* corresponds to the travelling rate between patches and *π*_ji_ denotes the rates of moving from origin *j* to destination *i*. The moving rates (*π*_ji_) were approximated by the radiation model, which showed an improved fit to data than the gravity model in the polio context [49–51]. We classified the infectious individual compartment into two classes: infectious individuals at patch *i* covered by the ES at day *t* (*I*_i,c,t_) and not covered by the ES (*I*_i,nc,t_). Infected individuals in the latent period (E) are randomly assigned to *I*_i,c,t_ or *I*_i,nc,t_ proportional to the patch-level ES population coverage (*p*_c_) and 1 - *p*_c_, respectively. However, assumptions on infectiousness did not differ between groups.

### AFP surveillance model

Patients with WPV1 develop AFP with a probability of 1/200, and we assumed those patients seek healthcare on the same day as the paralysis onset. The incubation period of developing AFP was assumed to be 16.5 days [52,53] and we prepared 6 compartments with transition rates of 0.329 days^-1^ to be aligned with the incubation period distribution [47].

To be correctly diagnosed and reported as AFP cases, patients should visit health care, be tested, and get a positive result. We assumed the probability of correctly reporting AFP cases followed the Binomial process considering the above three steps.

### Environmental surveillance model

ES was implemented in limited patches and its number was varied in each scenario. ES is assumed to cover a portion of the population (*p*_c_) in each patch. We chose a default *p*_c_ of 25% by considering the observed ES location and ES-covered population (Table S3-4 and Fig S8).

We modelled the detection process through ES as a binomial process. Let *n*_i,t_ be an index variable for sampling at location *i* at day *t* (taking 0 or 1), and *P*_ES,test_ be a test sensitivity given a collected sample contains a detectable level of virus. The number of polio-positive samples in the ES site at patch *i* at day *t* (*w*_i,t_) is given by

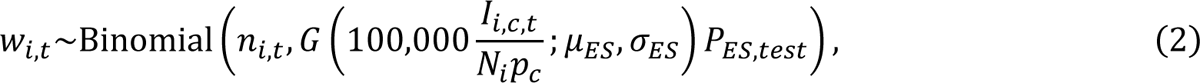

where *N*_i_ denotes the population size of all age groups in patch *i*, and *G* denotes the cumulative density function of the log-normal distribution with parameters of *μ*_ES_ and *σ*_ES_, which represents the ES sensitivity against the incidence rate of polio patients per 100,000 population. The ES sensitivity parameters were estimated based on the wastewater surveillance study on COVID-19 [40]. To ensure the consistent quality of ES in each sampling site for estimation, we used sampling sites with both positive and negative samples, where daily newly reported cases on the day with positive samples were consistently higher than on the day with negative samples. The estimated dose-relationship for the ES sensitivity can be seen in Fig S6 in S1 text.

### Sensitivity analysis and weighted minimum distance to ES-covered patches

We performed the sensitivity analyses of the basic reproduction number (*R*_0_), travelling rate between patches (α), sampling frequency and ES sensitivity for the IMP-POP/ES-POP scenario. For a patch-level ES population coverage (*p_c_*), sensitivity analysis was conducted for all 6 scenarios. Additionally, we conducted the sensitivity analysis under a single patch setting (i.e. simulations were performed without spatial structures). This analysis aimed to differentiate between the effects of parameters on the ES early detection ability attributable to spatial components and those stemming from model behaviours in a single patch (Fig S10 in S1 text).

Since our stochastic meta-population model requires a huge computational burden, we explored an alternative parsimonious assessment measurement for the ES early detection ability. Assuming the importation risk distributions and EIPs are known at a patch level, we calculated the average minimum distance to ES-covered patches weighted by importation risk and outbreak probability of 10 or more infections occurring (*d*_ave,w_). The outbreak probability was based on the branching process considering the effective reproduction number in patch *i* (*R*_e,i_) while the effective reproduction number was given by the product of *R*_0_ and EIP in patch *i* (Fig S9 in S1 text). We calculated the probability of 10 or more infections occurring given the effective reproduction number, *P*(*X* ≥ 10; *R*_*e,i*_), assuming the off-spring distribution follows the Poisson distribution [54–56]. Then, the average minimum distance *d*_ave,w_ is described as

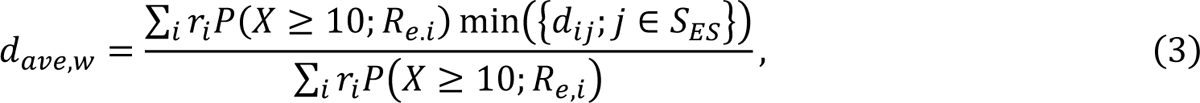

where *r*_i_ denotes the importation risk at patch *i* and *S*_ES_ denotes the set of patches covered by the ES. The detailed explanations can be found in S1 text.

## Data Availability

All the data and code are deposited on GitHub (https://github.com/toshiakiasakura/polio_environmental_surveillance).

## Acknowledgements

We thank Juliet R. C. Pulliam for insightful comments to improve result presentations and Kerrigan McCarthy for sharing wastewater plant-related information.

## Author contributions

Conceptualisation: TRA, KMO. Data curation: TRA. Formal analysis: TRA. Funding acquisition: TRA, KMO. Investigation: TRA. Methodology: TRA, KMO. Resources: TRA, KMO. Software: TRA. Supervision: KMO. Validation: TRA. Visualisation: TRA. Writing – original draft: TRA. Writing – review & editing: KMO.

## Supporting information

**S1 Text. Supplementary methods and results.**

**S1 Video. Simulated incremental implementation of environmental surveillance (ES) with the ‘Population size’-based ES site layout strategy (ES-POP).** ES sites were implemented in descending order of the population size of each patch. Blue squared areas represent patches covered by ES sites.

**S2 Video. Simulated incremental implementation of environmental surveillance (ES) with the ‘Land border crossing importation risk’-based ES site layout strategy (ES-LBC).** ES sites were first implemented in a patch with a high importation risk via land border crossing from Mozambique. Blue squared areas represent patches covered by ES sites.

## 1 Supplementary Methods

### 1.1 Data

We collated the population size data from the WorldPop [1] at 100m spatial resolution for 5 bin age categories and created population size data for children under 5 years old and for all ages in South Africa. We aggregated the population data into a 20km spatial resolution (precisely 21.31km x 19.74km), resulting in 3,193 patches. We removed patches with less than 100 children under 5 years old and finally obtained 1502 patches (Fig S1). Through this process, we removed 0.61% population of children under 5 years old. We kept patch spatial resolution constant for all patches so that ES-covered areas in each patch were fixed. We applied the same procedure to create the population data for children under 5 years old in Mozambique at the same resolution.

**Fig S1.**
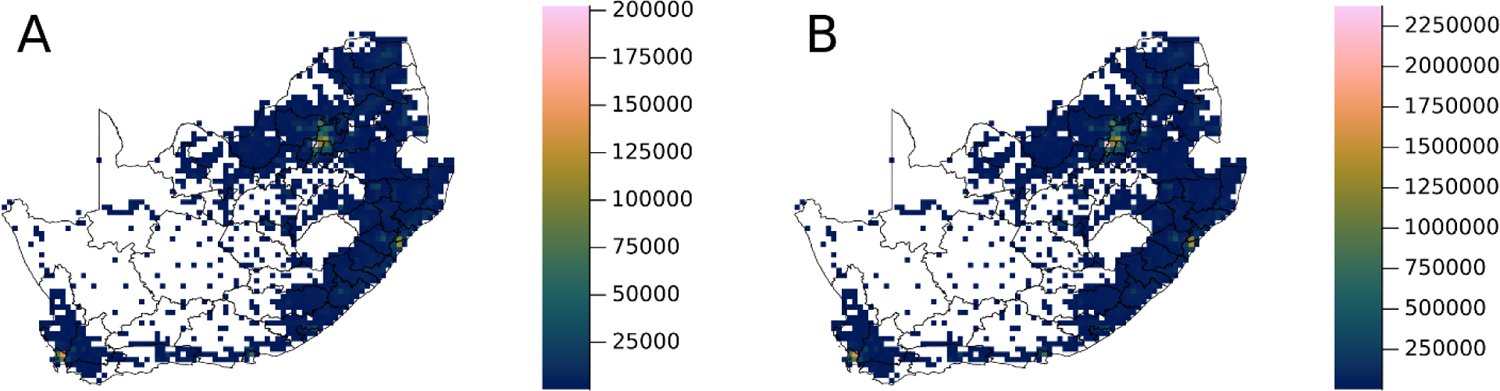
Heatmap of population size in South Africa. (A) Children under 5 years old and (B) All ages. Patches with <100 children under 5 years old were removed from the analysis.

We collated the district-level vaccination coverage from the Expanded Programme on Immunisation National Coverage Survey Report 2020 in South Africa [2]. We summarised the coverage of oral polio vaccine (OPV) and hexavalent vaccine (HEXA) including inactivated polio vaccine (IPV) for each district in Table S1 with the district-level effective immunisation proportions (EIPs), which were calculated using the equation described in the main text. We visualised the district-level EIPs in Fig S2.

**Table S1.** OPV coverage (%), HEXA coverage (%) and effective immunisation proportion (EIP, %) by districts in South Africa.

| District | Sample size | OPV0<sup>a</sup> | OPV1 | HEXA1<sup>b</sup> | HEXA2 | HEXA3 | HEXA4 | EIP<sup>c</sup> |
| --- | --- | --- | --- | --- | --- | --- | --- | --- |
| West Coast | 74 | 97.3 | 95.9 | 97.3 | 97.3 | 89.2 | 82.4 | 94.3 |
| Cape Winelands | 179 | 96.1 | 94.4 | 95.0 | 94.4 | 93.9 | 88.8 | 92.8 |
| Overberg | 143 | 97.2 | 94.4 | 94.4 | 94.4 | 96.5 | 84.6 | 94.3 |
| Eden | 121 | 96.7 | 88.4 | 95.9 | 95.0 | 90.9 | 89.3 | 93.3 |
| Central Karoo | 33 | 100.0 | 100.0 | 100.0 | 100.0 | 100.0 | 97.0 | 98.0 |
| Namakwa | 27 | 100.0 | 96.3 | 96.3 | 100.0 | 100.0 | 92.6 | 97.9 |
| Pixley ka Seme | 32 | 93.8 | 87.5 | 90.6 | 81.3 | 78.1 | 68.8 | 85.0 |
| Z F Mgcawu | 61 | 98.4 | 98.4 | 96.7 | 100.0 | 96.7 | 83.6 | 97.3 |
| Frances Baard | 200 | 99.0 | 98.0 | 99.0 | 99.5 | 98.5 | 90.5 | 97.3 |
| Cacadu | 348 | 97.1 | 95.4 | 96.0 | 96.3 | 94.8 | 87.6 | 94.1 |
| Amathole | 220 | 98.6 | 96.4 | 97.7 | 96.8 | 95.0 | 86.4 | 95.1 |
| Chris Hani | 79 | 98.7 | 92.4 | 92.4 | 97.5 | 93.7 | 87.3 | 95.0 |
| Joe Gqabi | 110 | 95.5 | 92.7 | 92.7 | 91.8 | 93.6 | 81.8 | 92.0 |
| O.R.Tambo | 372 | 95.4 | 90.3 | 86.6 | 87.9 | 84.4 | 76.3 | 85.6 |
| Xhariep | 81 | 97.5 | 95.1 | 96.3 | 92.6 | 93.8 | 76.5 | 93.8 |
| Lejweleputswa | 285 | 98.6 | 97.9 | 99.3 | 97.9 | 97.9 | 87.4 | 96.6 |
| Thabo Mofutsanyane | 50 | 100.0 | 96.0 | 98.0 | 98.0 | 98.0 | 86.0 | 95.8 |
| Fezile Dabi | 128 | 91.4 | 81.3 | 82.0 | 82.8 | 82.8 | 82.0 | 81.2 |
| Ugu | 187 | 97.3 | 96.8 | 97.9 | 98.4 | 96.3 | 88.8 | 96.1 |
| Umgungundlovu | 60 | 96.7 | 96.7 | 93.3 | 96.7 | 96.7 | 83.3 | 94.5 |
| Uthukela | 700 | 95.3 | 93.7 | 93.3 | 92.7 | 92.7 | 80.6 | 91.0 |
| Umzinyathi | 205 | 97.6 | 97.6 | 97.1 | 96.6 | 95.6 | 83.9 | 94.6 |
| Amajuba | 470 | 96.0 | 95.3 | 95.3 | 93.6 | 93.8 | 83.2 | 92.8 |
| Zululand | 399 | 96.5 | 95.5 | 95.0 | 95.2 | 93.7 | 77.9 | 92.7 |
| Umkhanyakude | 243 | 97.1 | 95.5 | 93.8 | 94.2 | 93.8 | 87.2 | 92.2 |
| Uthungulu | 986 | 92.4 | 92.3 | 92.7 | 91.7 | 91.3 | 82.8 | 90.3 |
| iLembe | 359 | 79.9 | 78.6 | 76.9 | 76.3 | 76.6 | 72.4 | 75.4 |
| Gert Sibande | 284 | 99.3 | 98.2 | 98.2 | 98.2 | 97.9 | 84.9 | 95.9 |
| Nkangala | 328 | 90.2 | 87.5 | 88.7 | 88.4 | 86.9 | 78.0 | 86.5 |
| Ehlanzeni | 396 | 93.2 | 92.9 | 89.9 | 89.9 | 89.6 | 81.1 | 87.9 |
| Mopani | 680 | 93.5 | 91.5 | 91.9 | 92.1 | 90.4 | 81.6 | 89.9 |
| Vhembe | 589 | 93.0 | 92.5 | 92.2 | 91.9 | 90.8 | 82.0 | 90.0 |
| Capricorn | 493 | 87.4 | 85.8 | 86.0 | 86.6 | 85.8 | 71.6 | 84.4 |
| Waterberg | 111 | 96.4 | 95.5 | 96.4 | 94.6 | 92.8 | 73.9 | 93.1 |
| Bojanala | 299 | 94.3 | 91.6 | 93.0 | 91.3 | 90.6 | 83.3 | 90.3 |
| Ngaka Modiri Molema | 484 | 97.1 | 96.7 | 94.8 | 95.2 | 93.2 | 82.0 | 92.8 |
| Dr Ruth Segomotsi Mompati | 106 | 97.2 | 97.2 | 97.2 | 95.3 | 95.3 | 84.9 | 94.4 |
| Dr Kenneth Kaunda | 210 | 95.7 | 94.8 | 94.8 | 93.3 | 91.9 | 83.8 | 92.1 |
| Sedibeng | 514 | 94.6 | 92.6 | 94.2 | 93.8 | 93.0 | 84.0 | 91.9 |
| Sisonke | 597 | 96.0 | 95.1 | 94.6 | 94.1 | 94.5 | 87.1 | 92.8 |
| Alfred Nzo | 271 | 98.2 | 95.9 | 96.7 | 96.7 | 95.6 | 83.4 | 94.4 |
| John Taolo Gaetsewe | 5 | 100.0 | 100.0 | 100.0 | 100.0 | 100.0 | 100.0 | 98.1 |
| Sekhukhune | 345 | 92.8 | 89.6 | 91.6 | 91.9 | 90.7 | 82.6 | 89.8 |
| West Rand | 382 | 97.4 | 96.9 | 97.4 | 96.1 | 95.8 | 90.1 | 94.9 |
| Buffalo City | 211 | 98.6 | 95.3 | 95.3 | 95.3 | 94.8 | 86.3 | 93.2 |
| City of Cape Town | 101 | 97.0 | 98.0 | 97.0 | 97.0 | 94.1 | 90.1 | 94.7 |
| Ekurhuleni | 890 | 97.1 | 96.1 | 96.6 | 95.5 | 94.9 | 86.9 | 94.1 |
| eThekweni | 887 | 94.0 | 92.6 | 92.4 | 91.7 | 91.8 | 83.8 | 90.3 |
| City of Johannesburg | 1710 | 97.0 | 96.4 | 95.6 | 95.3 | 94.8 | 87.8 | 93.4 |
| Mangaung | 187 | 98.9 | 97.9 | 97.3 | 98.4 | 97.9 | 87.7 | 96.2 |
| Nelson Mandela Bay | 326 | 94.5 | 93.3 | 93.9 | 92.9 | 92.0 | 86.5 | 91.5 |
| City of Tshwane | 621 | 98.9 | 96.3 | 96.5 | 95.3 | 95.8 | 86.5 | 94.5 |

<sup>a</sup> OPV: Oral polio vaccine
<sup>b</sup> HXA: Hexavalent vaccine including diphtheria, tetanus, acellular pertussis, Haemophilus influenzae type b, inactivated polio vaccine (IPV), and hepatitis B vaccines.
<sup>c</sup> EIP: Effective immunisation proportion.

**Fig S2.**
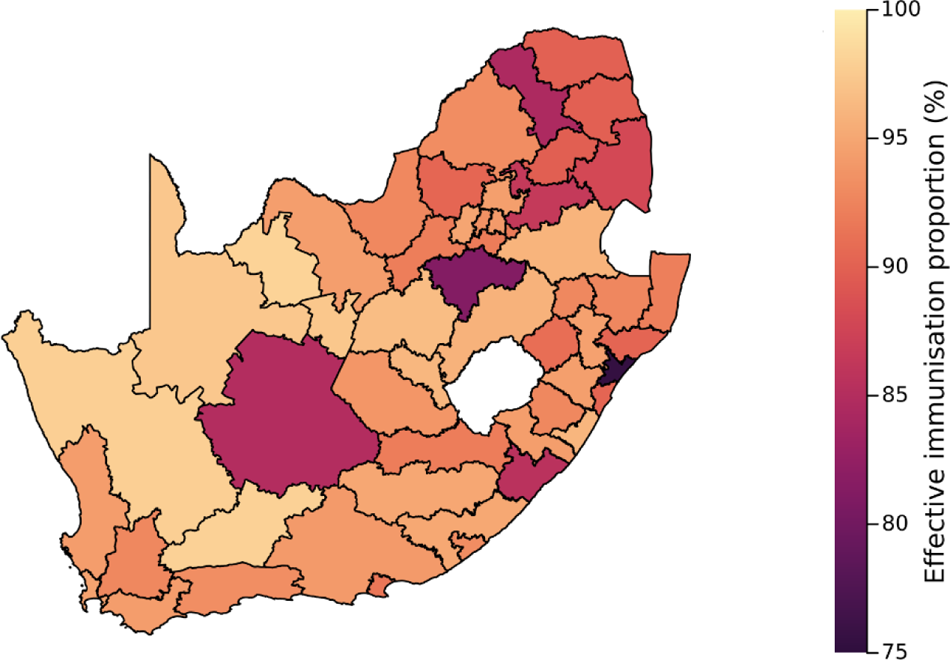
Estimated effective immunisation proportion by districts in South Africa, 2020.

### 1.2 Ethical statement

This study was assessed by the Research Governance & Integrity Office at the London School of Hygiene & Tropical Medicine as not requiring ethical approval due to its study design as a secondary data analysis on 9 May 2023 (MSc ethics reference number: 29004). The location and population size covered by each wastewater plant were provided by the National Institute of Communicable Diseases (NICD) in South Africa. All other data are publicly available.

### 1.3 Modelling framework

To assess the early detection ability of environmental surveillance (ES) over acute flaccid paralysis (AFP) surveillance, we constructed the stochastic spatiotemporal model among unimmunised children under 5 years old considering the detection process through the AFP surveillance and ES (Fig S3). We utilised South Africa as a case study of a non-endemic country and assumed a single introduction of a patient with wild poliovirus serotype 1 (WPV1). Following other modelling studies on polio [3–5], our model comprises three components: transmission model, AFP surveillance model and ES model (Fig S3), which will be explained in later sections. We used the detection patterns and first detection timing by ES and AFP surveillance as outcomes to quantify the early detection ability while we varied the number of ES-covered patches. We prepared three importation risk distributions and two ES site layout strategies, totalling six scenarios, to account for limited knowledge of importation risks (Fig 1).

**Fig S3.**
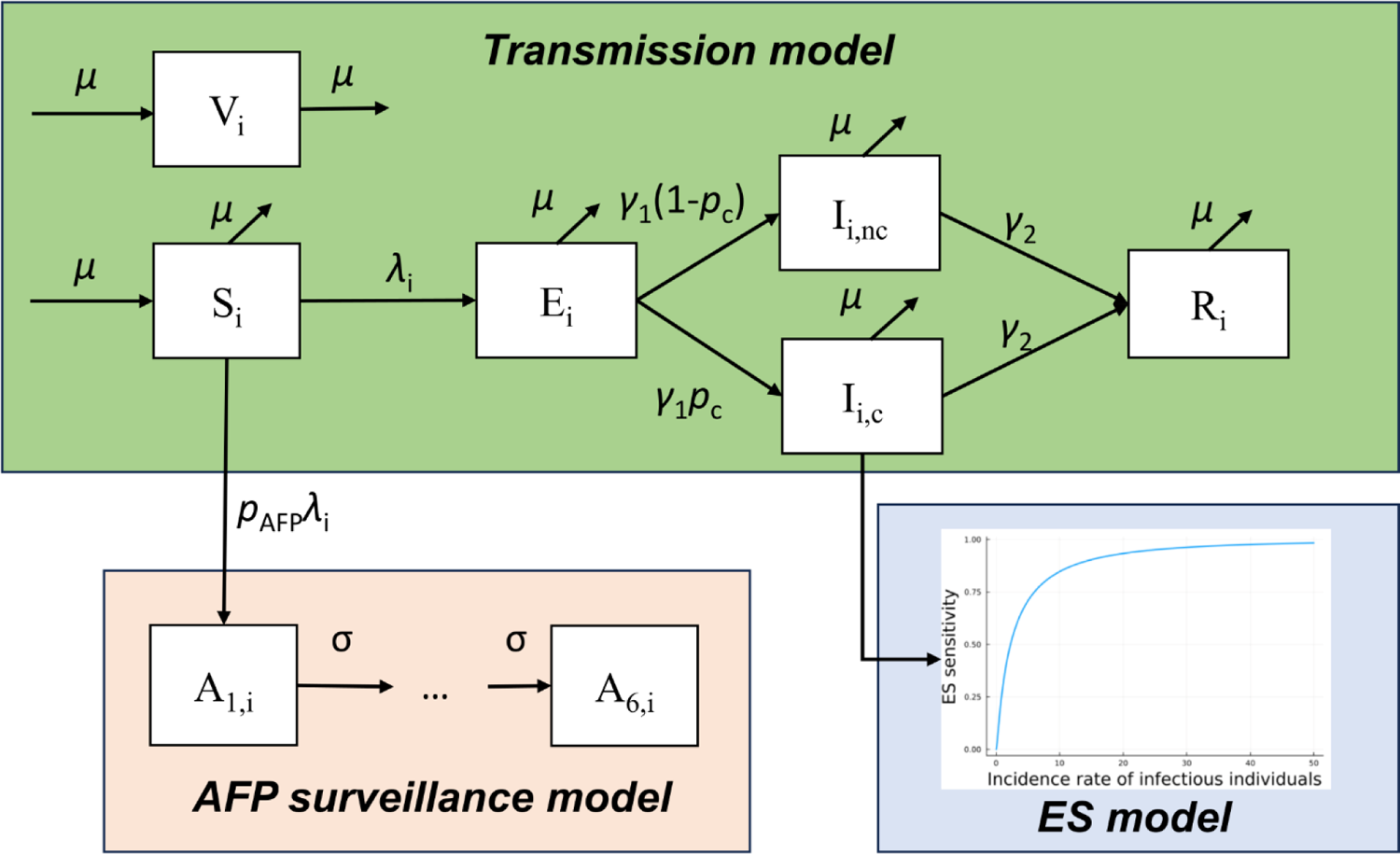
Schematic representation of our model, comprising of three parts: transmission model, AFP surveillance model and ES model. Each compartment is defined as follows: S, susceptible population; E, latent population; I, infectious population; R, recovered population; V, immunised population; and A_1_ through A6, population who are developing AFP. ES sensitivity is dependent on the incidence rate of infectious polio patients. The subscript of *i* corresponds to the location. Abbreviations: ES, environmental surveillance; AFP, acute flaccid paralysis.

#### 1.3.1 Transmission model

We employed the stochastic spatiotemporal SEIR model to describe the transmission dynamics. The SEIR model simulates the process in which susceptible individuals (S) get infected, passing through a latent period (E) and infectious period (I), and then recover from infection (R). We only considered unimmunised children under 5 years old since most reported patients with AFP were within this age group [6]. We assumed no waning of immunity from vaccination or natural infection. Considering our simulation length (i.e. 3 years), we introduced the birth and removal rate (*μ*) to reflect the population dynamics. We classified infectious compartments into the one covered by the ES (*I*_c_) and the one not covered by the ES (*I*_nc_) but assumptions on infectiousness did not differ between groups.

The force of infection at patch *i* at day *t* (*λ*_i,t_) corresponds to the rate at which susceptible individuals at patch *i* at day *t* get infected and is described by

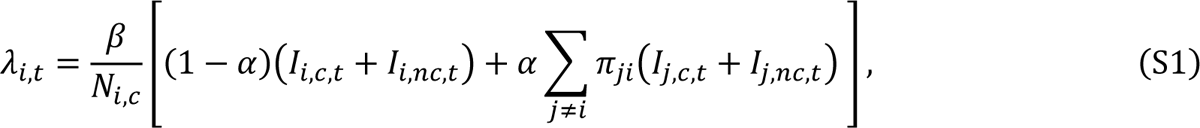

where *β* corresponds to the transmission rate, and *α* corresponds to the travelling rate between patches, which represents the proportion of travellers per day from one patch. The population aged under 5 years old at patch *i* is denoted as *N*_i,c_. It is noted that immunised populations (*V*_i_) contributed to herd immunity and its population size was given by *N*_i,c_ multiplied by the EIP at patch *i* (EIP_i_). Then, the initial susceptible population at patch *i* (*S*_i_) was given by *N*_i,c_ (1-EIP_i_), and the effective reproduction number at patch *i* at the beginning of the simulation (*R*_e,i_) was given by the product of the basic reproduction number (*R*_0_=β/γ_2_) and the EIP at patch *i*.

The second term of Equation S1 corresponds to the hazard from other patches, considering the travelling rate (α) and moving rates of travellers from origin *j* to destination *i* among all travellers moving from patch *j* (*π*_ji_). We adopted the frequency-dependent model for between-patch transmission, which was theoretically investigated [7,8], while another modelling study on poliovirus employed density-dependent transmission [5]. We used the radiation model to approximate moving rates (*π*_ji_):

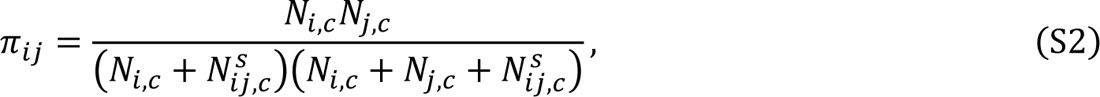

where *N*^*s*^_ij,c_ denotes the aggregated population aged under 5 years old within the circular area with a radius extending from patch *i* to patch *j*, centred at patch *i* but excluding the source and destination population. The histogram and heatmap of moving rates (*π*_ij_) of the top 3 populous patches are shown in Fig S4. The previous study suggested in the context of polio disease, the radiation model not only outperformed the gravity model [9–11], but also excelled when compared to the gravity model calibrated to mobile phone data [9].

We adopted the discrete Markov process with daily time steps for our stochastic simulations. Transition events, effects and sampling ways of new states were summarised in Table S2.

**Fig S4.**
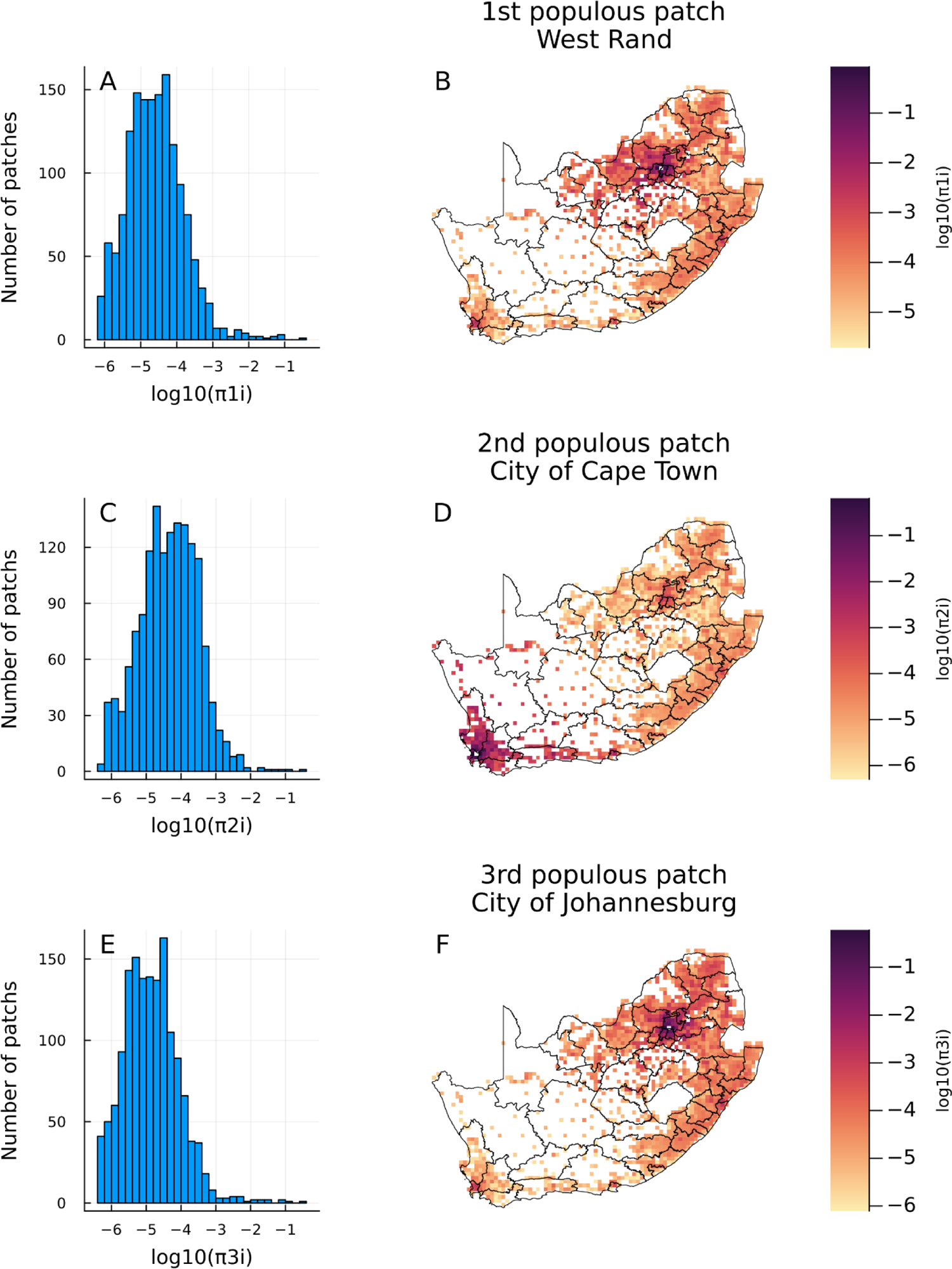
Moving rates from origin *i* to destination *j* (*π*_ij_) approximated by the radiation model for the three most populous patches. (A, C, E) Histogram of *π*_ij_ and (B, D, E) heatmap of *π*_ij_ with colour bar values on a log10 scale for (A, B) West Rand, (C, D) City of Cape Town and (E, F) City of Johannesburg.

**Table S2.** Discrete Markov process for each time step.

| Event (patch $i$) | Effect | Sampling way |
| --- | --- | --- |
| Birth of susceptible individuals | $(S_{i,t+1}) \leftarrow (S_{i,t} + Z_{i,t}^{(N_{i,u})})$ | $Z_{i,t}^{(N_{i,u})} \sim \text{Binomial}(N_{i,u}, 1 - e^{-\mu})$ |
| Removal of susceptible individuals because of age eligibility | $(S_{i,t+1}) \leftarrow (S_{i,t} - Z_{i,t}^{(S_{i,t})})$ | $Z_{i,t}^{(S_{i,t})} \sim \text{Binomial}(S_{i,t}, 1 - e^{-\mu})$ |
| Infection of susceptible individuals | $(S_{i,t+1}, E_{i,t+1}) \leftarrow (S_{i,t} - Z_{i,t}^{(S_{i,t}, E_{i,t})}, E_{i,t} + Z_{i,t}^{(S_{i,t}, E_{i,t})})$ | $Z_{i,t}^{(S_{i,t}, E_{i,t})} \sim \text{Binomial}(S_{i,t}, 1 - e^{-\lambda_{i,t}})$ |
| New individuals who will develop AFP | $(A_{1,i,t+1}) \leftarrow (A_{1,i,t} + Z_{i,t}^{(A_{1,i,t})})$ | $Z_{i,t}^{(A_{1,i,t})} \sim \text{Binomial}(Z_{i,t}^{(S_{i,t}, E_{i,t})}, p_{AFP})$ |
| Removal of latent individuals because of age eligibility | $(E_{i,t+1}) \leftarrow (E_{i,t} - Z_{i,t}^{(E_{i,t})})$ | $Z_{i,t}^{(E_{i,t})} \sim \text{Binomial}(E_{i,t}, 1 - e^{-\mu})$ |
| Becoming infectious | $(E_{i,t+1}, I_{i,t+1}) \leftarrow (E_{i,t} - Z_{i,t}^{(E_{i,t}, I_{i,t})}, I_{i,t} + Z_{i,t}^{(E_{i,t}, I_{i,t})})$ | $Z_{i,t}^{(E_{i,t}, I_{i,t})} \sim \text{Binomial}(E_{i,t}, 1 - e^{-\gamma_1})$ |
| Removal of infected individuals because of age eligibility | $(I_{i,t+1}) \leftarrow (I_{i,t} - Z_{i,t}^{(I_{i,t})})$ | $Z_{i,t}^{(I_{i,t})} \sim \text{Binomial}(I_{i,t}, 1 - e^{-\mu})$ |
| Recovery from infection | $(I_{i,t+1}, R_{i,t+1}) \leftarrow (I_{i,t} - Z_{i,t}^{(I_{i,t}, R_{i,t})}, R_{i,t} + Z_{i,t}^{(I_{i,t}, R_{i,t})})$ | $Z_{i,t}^{(I_{i,t}, R_{i,t})} \sim \text{Binomial}(I_{i,t}, 1 - e^{-\gamma_2})$ |
| Removal of recovered individuals because of age eligibility | $(R_{i,t+1}) \leftarrow (R_{i,t} - Z_{i,t}^{(R_{i,t})})$ | $Z_{i,t}^{(R_{i,t})} \sim \text{Binomial}(R_{i,t}, 1 - e^{-\mu})$ |
| Progression of the incubation period of AFP from stage $k$ to $k+1$ | $(A_{k,i,t+1}, A_{k+1,i,t+1}) \leftarrow (A_{k,i,t} - Z_{i,t}^{(A_{k,i,t}, A_{k+1,i,t})}, A_{k+1,i,t} + Z_{i,t}^{(A_{k,i,t}, A_{k+1,i,t})})$ | $Z_{i,t}^{(A_{k,i,t}, A_{k+1,i,t})} \sim \text{Binomial}(A_{k,i,t}, 1 - e^{-\sigma})$ |

<sup>a</sup> $N_{i,u}$ denotes unvaccinated individuals at patch $i$ , calculated as the product of population aged under 5 years old and effective immunisation proportion at patch $i$ .

We estimated the infectious period (1/γ_2_) using the proportion of infected individuals shedding any amount of virus over time, which was provided by expert opinions [12]. Formally, the time course of infectiousness can be obtained by the product of the proportion of individuals shedding any amount of virus and non-linear transformation of the amount of virus shedding given a person excretes the virus each day since infection [12]. Since we do not know the functional form for the latter data, we regarded the first one as the infectiousness for simplicity.

Since the compartment model implicitly assumes the transition process from one compartment to another follows the exponential distribution, the distribution of infectiousness over time can be described as the convolution of two exponential distributions:

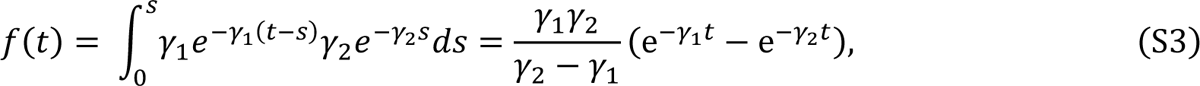

where the reciprocal, 1/γ_1_, corresponds to the mean incubation period and is assumed to be 4 day, and 1/γ_2_ corresponds to the mean infectious period. We minimised the Kullback-Leibler divergence between the observed distribution and *f(t)*, estimating 1/γ_2_ to be 15.02 days. The fitted curve is shown in Fig S5.

**Fig S5.**
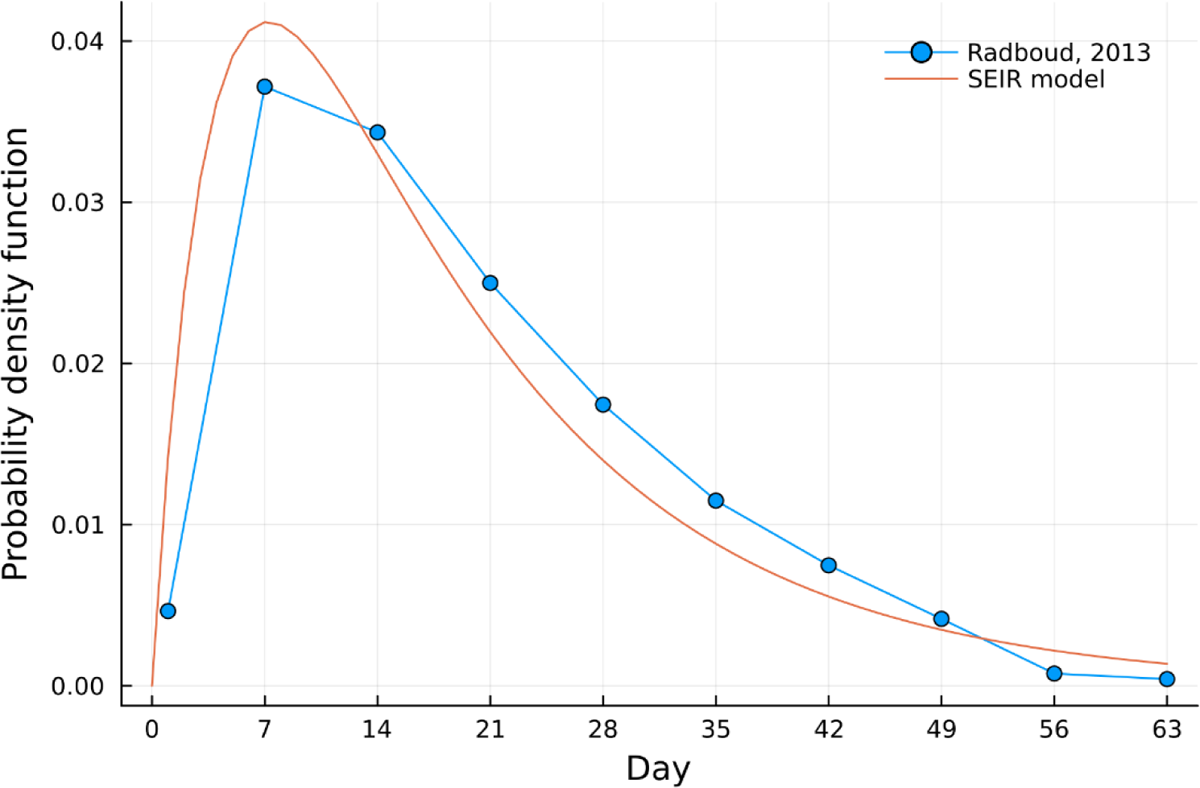
The proportion of individuals excreting the poliovirus regardless of the amount of virus shedding over time, which was scaled to be one for the probability density function and the fitted probability density function.

#### 1.3.2 AFP surveillance model

We assumed the probability of developing AFP given infection with poliovirus to be 1/200, which was denoted as *p*_AFP_. Patients developing AFP are correctly reported if they successfully undergo the following process: seeking healthcare, being tested, and getting a positive result. We denoted *P*_H_, *P*_AFP, sample_, and *P*_AFP, test_ as the probability of each process. Then, the number of reported patients with AFP at patch *i* at time *t*, O_i,t_, was described as

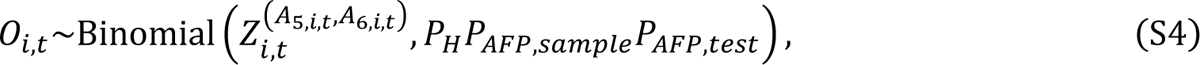

where the number of individuals who newly develop AFP at patch *i* at day *t* is denoted as 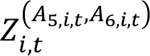

The incubation period of developing AFP was assumed to be 16.5 days [13,14] and we prepared 6 compartments with transition rates of 0.329 days^-1^ to be aligned with the incubation period distribution [5]. The number of compartments and transition rates were estimated in the previous study [5] by fitting an Erlang distribution to 36 independent data intervals from poliovirus exposure to the onset of AFP. In the present study, we did not consider a delay in medical consultations, conducting tests and notifications to authorities. These processes usually take time [15] and should be accounted for when results are interpreted.

#### 1.3.3 Environmental surveillance model

There are limited empirical data to compare the incidence of poliovirus against the probability of a positive ES sample, which is a required parameter in this analysis. Instead, we opted to assume that ES sensitivity for poliovirus was comparable to that observed for COVID-19, and used the wastewater surveillance data in the US, published by Fuqing Wu et al. [16]. The data for COVID-19 is more amenable to analysis because of the intensity of clinical testing for SARS-CoV-2 and the (comparatively) low proportion of asymptomatic infections. To ensure the consistent quality of ES in each sampling site for estimation, we used sampling sites with both positive and negative samples, where daily newly reported cases on the day with positive samples were consistently higher than on the day with negative samples. Finally, 27 out of 353 sampling sites were eligible for the estimation.

We employed the log-normal distribution to estimate the dose-response curve for the ES sensitivity against the incidence rate of COVID-19 (thus also of poliovirus infection). Let *x*_i,t_ be the number of newly reported COVID-19 cases at sampling site *i* on day *t*, *y*_i,t_ be the indicator variable for an observed positive sample at sampling site *i* on day *t,* and *p*_i,t_ be the probability of detecting a positive sample at site *i* on day *t*. We minimized the following likelihood function to estimate the parameters of the lognormal distribution (*μ*_ES_ and *σ*_ES_):

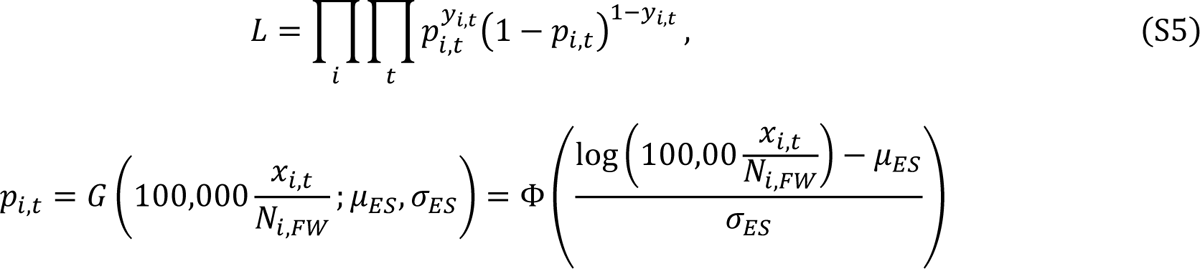

where *G* denotes the cumulative density function of the log-normal distribution, *N*_i, FW_ denotes the population in the state with sampling site *i,* which was given in Fuqing Wu et al. [16], and Φ corresponds to the cumulative density function of the standard normal distribution. We obtained 0.818 for *μ*_ES_ and 1.45 for *σ*_ES_.

The estimated parameters here will be inappropriate in terms of differences in the amount and length of virus shedding between SARS-CoV-2 and poliovirus in addition to differences in diagnostics for the detection of virus. In the main text, we conducted the sensitivity analysis ranging the ES sensitivity from 10 times higher to 10 times lower. We multiplied 1/k for previous data to obtain parameters for k times higher ES sensitivity. The dose-response curves for those ES sensitivity parameters are shown in Fig S6. *σ*_ES_ was estimated to be constant across different ES sensitivity assumptions and *μ*_ES_ was estimated to be 3.121, 1.917, −0.281 and −1.485 for 10 times lower, 3 times lower, 3 times higher, and 10 times higher ES sensitivity, respectively.

**Fig S6.**
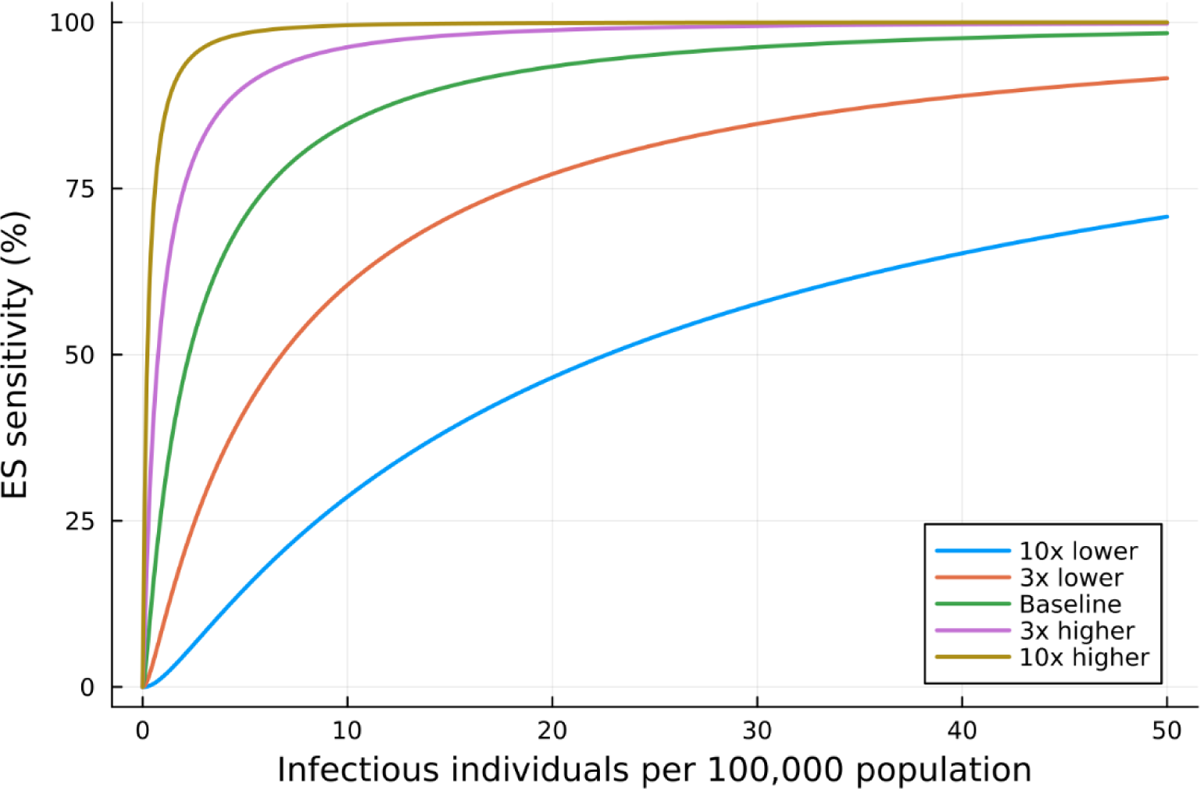
Dose-response curve for the ES sensitivity parameter against the number of infectious individuals per 100,000 population in a single patch.

We modelled the detection process through ES as the binomial process in our simulation study. Let *n*_i,t_ be the indicator variable for conducting environmental sampling at patch *i* at day *t* (e.g. given monthly sampling, *n*_i,t_ takes 1 on a single day, and takes 0 for the remaining days within a month), and *P*_ES, test_ be the test sensitivity given a collected sample contains a detectable level of virus. The number of polio-positive samples in wastewater (*w*_i,t_) is given by

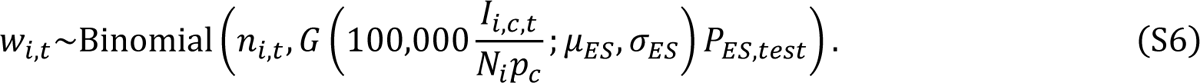

It is noted that in each patch, *p*_c_% of the population was assumed to be covered by the ES, and we separately considered the infectious individuals covered by ES (*I*_i,c,t_) or not (*I*_i,nc,t_) in the compartment model. Here *N*_i_ denotes the population of all ages at patch *i* (not limited to children under 5 years old).

#### 1.3.4 Importation risk distributions

Importation risk at patch *i* refers to the probability that WPV1 is introduced to that specific patch and importation risk distribution refers to the distribution of importation risk across patches. We considered three importation risk distributions, which are denoted as IMP-POP, IMP-AIR, and IMP-LBC.

The IMP-POP refers to the ‘Population size’-based importation risk distribution and assumes importation risk is proportional to the population size of each patch. Let *r*_i_ be the importation risk at patch *I* for the IMP-POP scenario and be described as

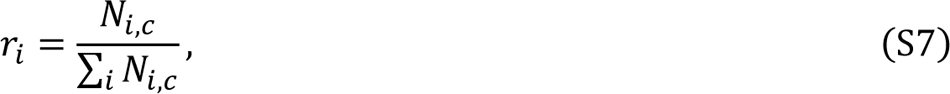

where *N*_i,c_ denotes the population size of children aged under 5 years old at patch *i*.

The IMP-AIR refers to the ‘International airport’-based importation risk distribution and assumes importation risk is proportional to international inbound travel volume in 2019 and further considers mobilisation from airports, which was approximated by the radiation model given in Equation S2.

Without considering mobilisation from airports, importation risks would be too confined to a few patches, which are sometimes very rural areas and we did not think that importation risk distribution is a realistic one. South Africa holds three international airports and their international inbound travel volumes in 2019 were 4,342,611 for O.R. Tambo International Airport, 1,156,996 for Cape Town International Airport, and 188,243 for King Shaka International Airport (Fig 1D).

We denote *l*_1_*, l*_2_ and *l*_3_ as the location of a patch with O.R. Tambo International Airport, Cape Town International Airport, and King Shaka International Airport, respectively. Let *w*_li_ be the proportion of the inbound travel volume at patch *l*_i_ with international airport *i*. The importation risk at patch *i* for the IMP-AIR is described as the average of moving rates from three international airports weighted by their inbound travel volumes:

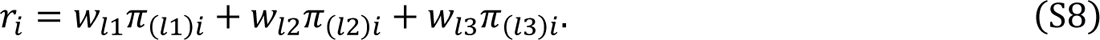

The IMP-LBC refers to the ‘Land border crossing’-based importation risk distribution and assumes importation risk is proportional to travelling volume from Mozambique. Here, we assumed poliovirus was circulating in Mozambique and importation from Mozambique to South Africa happened via land border crossing. Since mobilisation volume data between Mozambique and South Africa is not available, we approximated those mobilisations with the radiation model.

The probability of the presence of poliovirus in patch *j* in Mozambique was assumed to be proportional to the population size of children under 5 years old, and the movement from patch *j* in Mozambique to patch *i* in South Africa is given by the radiation model between the two countries. The importation risk at patch *i* in South Africa for the IMP-LBC is described as

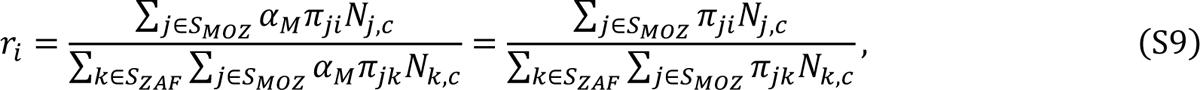

where *S*_MOZ_ and *S*_ZAF_ represent the set of patches in Mozambique and South Africa, respectively. The travelling rate from Mozambique to South Africa (*α*_M_) would be different from the travelling rate between patches in South Africa (*α*) but is cancelled out in Equation S9.

The importation risk distributions explained above are visualised in Fig S7.

**Fig S7.**
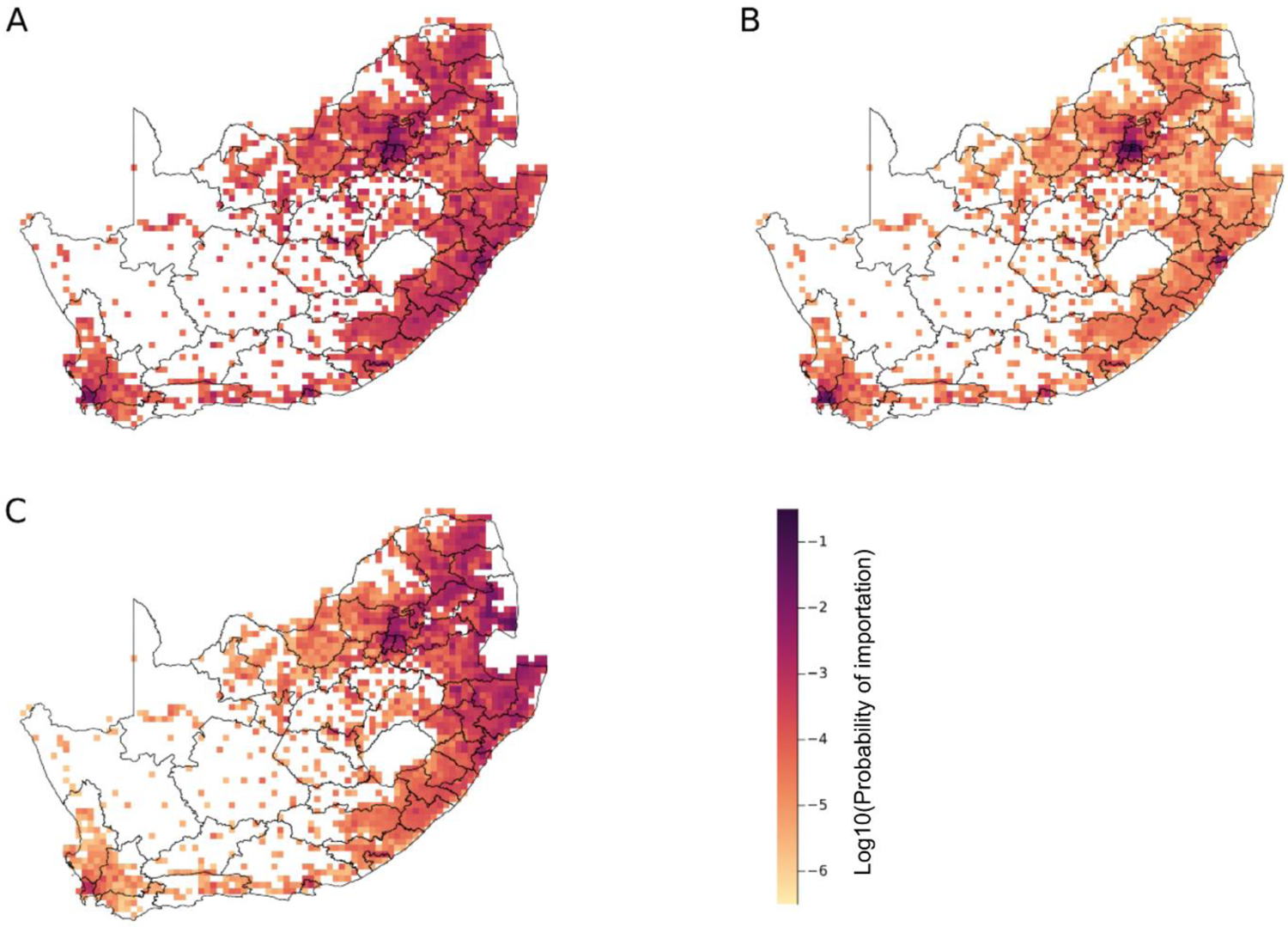
Importation risk distributions in a log10 scale (A) for IMP-POP, (B) IMP-AIR, and (C) IMP-LBC.

#### 1.3.5 ES site layout strategies and a patch-level ES population coverage (*p*c)

ES site layout strategy determines a sequence of which patches are covered by the ES when we increase the number of ES-covered patches. We considered two ES site layout strategies: ES-POP and ES-LBC. The ES-POP refers to the ‘Population size’-based ES site layout strategy, and assumes the ES is implemented in the descending order of population size of each path. Hence, this strategy can cover the largest population size by the ES given the same number of ES-covered patches.

The ES-LBC refers to the ‘Land border crossing importation risk’-based ES site layout strategy, and assumes the ES is first implemented in a patch with a high importation risk via land border crossing from Mozambique. The importation risk from Mozambique is given as the same in Equation S9. The motivation to prepare the ES-LBC is to quantify the effectiveness of the strategic positioning of the ES sites against the high importation risks in rural areas. The animations of the incremental ES implementation from a single patch to all patches were prepared for both ES-POP and ES-LBC in gif-files (S1-2 Video). It is noted that the maximum national ES population coverage was set at 25% since we assumed the ES-population coverage in each patch (called a patch-level ES population coverage, *p*_c_) to be 25%, which reason is explained below.

We aimed for our simulated ES site layout under the ES-POP to be closely aligned with the observed one. The ES site is generally prioritised to be placed in populous areas in the real-world setting. To quantify the similarity between simulated and observed ES layouts in South Africa, we first calculated the observed national and district-level ES population coverage (Table S3). Then, we varied a patch-level ES population coverage (*p*_c_) fixing the simulated national ES population coverage to be the same as the observed one, and descriptively compared the simulated number of districts with ES sites and the corresponding district-level ES population coverage with the observed ones (Table S4).

We calculated the observed national and district-level ES population coverage using the location and ES-covered population of each wastewater plant, which was provided by the National Institute of Communicable Diseases in South Africa as of 27 November 2023. The ES-covered population was missing for Rooiwai Eastern and Daspoort wastewater plants and we imputed the median ES-covered population size of 350,000. We finally estimated the observed national ES population coverage in South Africa to be 11.3% (Table S3). The median and mean of the district-level ES population coverage among districts with ES sites were 30.6% and 22.5%, respectively.

We simulated the ES layout varying the patch-level ES population coverage (*p*_c_) while fixing the simulated national ES population coverage to be the observed one of 11.3% (Table S4). When we set *p*_c_ at 100%, the ES sites were too concentrated and simulated district-level ES population coverages were much higher than observed ones. We chose *p*_c_ to be 25% for the main analysis considering the dispersion of ES sites and district-level ES population coverages (Table S4 and Fig S8).

**Table S3.** Approximated ES-covered population size in districts with ES sites in South Africa as of 27 November 2023.

| Province | District | District-level population size | Wastewater plant name | ES-covered population size | District-level ES population coverage (%) |
| --- | --- | --- | --- | --- | --- |
| Eastern Cape | Buffalo City | 760840 | East Bank | 141000 | 22.5 |
|  |  |  | Gonubie | 30400 |  |
| Free State | Mangaung | 826621 | Bloemspruit | 350000 | 66.5 |
|  |  |  | Sterkwater | 200000 |  |
| Gauteng | City of Johannesburg | 5540727 | Northern | 1200000 | 46.0 |
|  |  |  | Goudkoppies | 500000 |  |
|  |  |  | Bushkoppies | 850000 |  |
|  | City of Tshwane | 3627986 | Rooiwal Eastern | NA | NA |
|  |  |  | Daspoort | NA |  |
|  | Ekurhuleni | 3739653 | Hartebeesfontein | 100000 | 10.7 |
|  |  |  | Olifantsfontein | 100000 |  |
|  |  |  | Vlakplaats | 200000 |  |
| KwaZulu Natal | eThekweni | 3587907 | Central | 350000 | 18.6 |
|  |  |  | Northern | 316425 |  |
| North West | Bojanala | 1786678 | Rustenburg | 509000 | 28.5 |
| Western Cape | City of Cape Town | 4436413 | Borchard's Quarry | 380000 | 18.9 |
|  |  |  | Zandvliet | 460000 |  |

**Table S4.** Observed and simulated district-level ES population coverage for the ES-POP varying the patch-level ES population coverages (*p*_c_) given the simulated national ES population coverage was fixed at the observed one (11.3%).

| Province | District | Observed district-level ES population coverage (%) | Simulated district-level ES population coverage (%) |  |  |
| --- | --- | --- | --- | --- | --- |
| | | | $p_c=1.0^a$ | $p_c=0.5^a$ | $p_c=0.25^a$ |
| Eastern Cape | Buffalo City | 22.5 |  |  | 19.6 |
|  | Nelson Mandela Bay | 0.0 |  | 36.0 | 18.0 |
| Free State | Mangaung | 66.5 |  |  | 15.2 |
|  | Thabo Mofutsanyane | 0.0 |  |  | 9.7 |
| Gauteng | Ekurhuleni | 10.7 |  | 17.5 | 25.0 |
|  | City of Johannesburg | 46.0 | 37.1 | 50.0 | 25.0 |
|  | City of Tshwane | 19.3 |  | 12.4 | 21.2 |
|  | West Rand | 0.0 | 56.8 | 28.4 | 23.6 |
|  | Sedibeng | 0.0 |  |  | 17.5 |
| Kwazulu Natal | eThekweni | 18.6 | 64.2 | 32.1 | 22.2 |
|  | iLembe | 0.0 |  | 31.8 | 19.3 |
|  | Umgungundlovu | 0.0 |  |  | 8.8 |
|  | Amajuba | 0.0 |  |  | 9.1 |
| Western Cape | City of Cape Town | 18.9 | 36.7 | 40.7 | 22.8 |
|  | Cape Winelands | 0.0 |  |  | 12.5 |
| Limpopo | Waterberg | 0.0 |  |  | 8.6 |
|  | Capricorn | 0.0 |  |  | 7.9 |
| Mpumalanga | Ehlanzeni | 0.0 |  |  | 7.0 |
| North West | Bojanala | 28.5 |  |  | 11.6 |
| Northern Cape | NA | NA |  |  |  |

<sup>a</sup> Blank in those columns represents no ES-covered patches.

**Fig S8.**
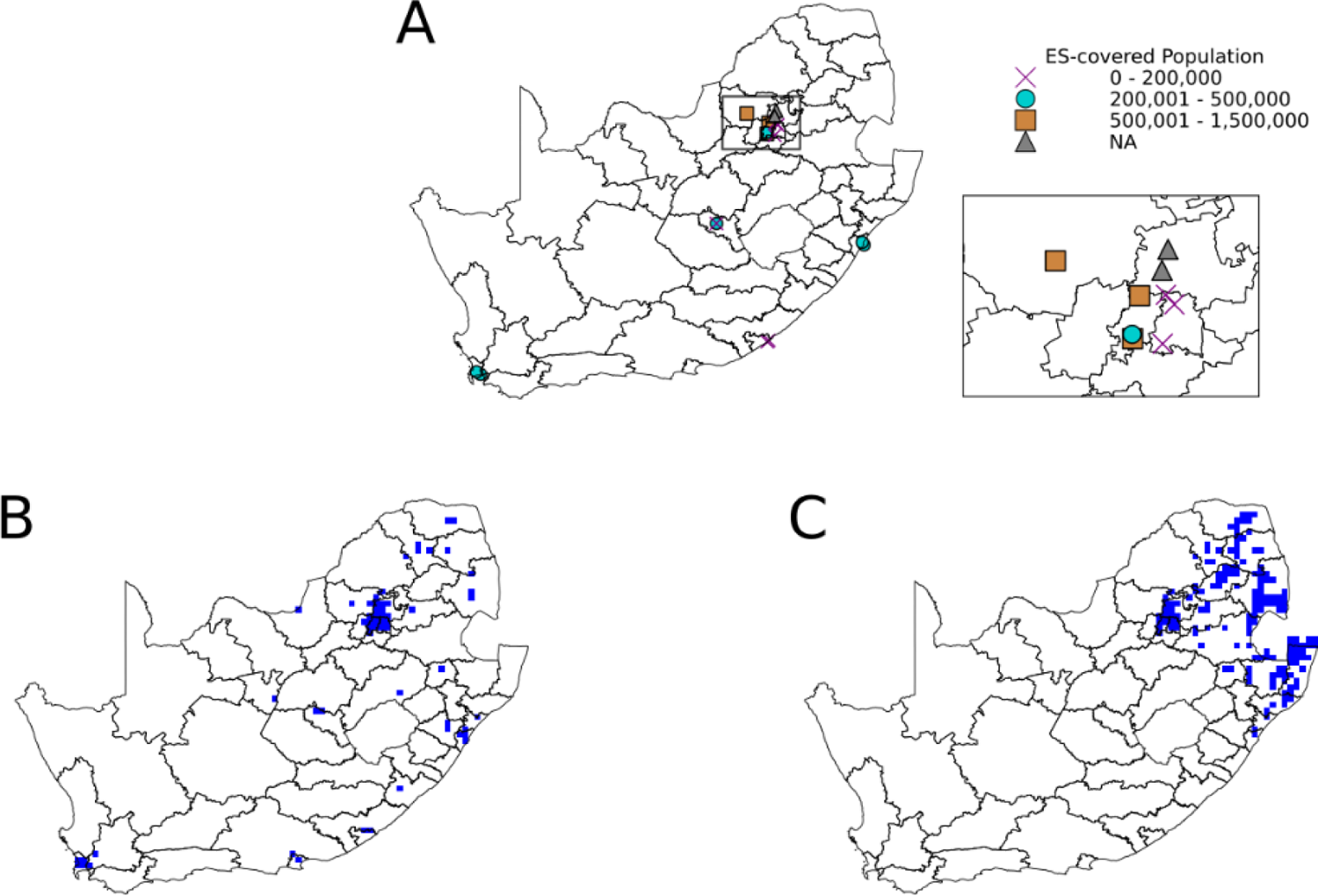
Environmental surveillance (ES) location maps. (A) Observed 17 ES sites in South Africa. Data points represent the location of observed ES sites, with each marker style indicating a specific range of ES-covered population sizes. The inset displays the expanded view of the areas with a high density of ES sites. (B, C) Simulated ES site layout when the simulated national ES population coverage was matched with the observed coverage given the patch-level ES population coverage was set at 25% (B) for ES-POP and (C) for ES-LBC. Blue squared areas represent patches with simulated ES sites. The number of ES-covered patches was 58 for ES-POP and 154 for ES-LBC.

#### 1.3.6 Model parameter specification

Parameter values used in our simulations are summarised in Table S5. We explained the rationale for the choice of several parameters.

We chose the basic reproduction number (*R*_0_) to be 14 based on the estimation for developing countries by Fine et al.1999, which used age-stratified seroprevalence data [17]. This estimated value would be higher considering the current hygiene situation in South Africa. In our transmission model, the population who were successfully vaccinated and immunised were removed from the dynamics. We define the transmissibility accounting for the initial susceptible population as the effective reproduction number at the beginning of the simulation for South Africa, *R*_e_(0). We calculated *R*_e_(0) by the product of *R*_0_ and EIP in each district and averaged them across districts weighted by the population size of each district, and we obtained 1.13 for *R*_e_(0).

Regarding the AFP surveillance-related parameters, the stool adequacy rate refers to two stool specimens of sufficient quantity for laboratory analysis, collected at least 24 hours apart, within 14 days after the onset of paralysis. The stool adequacy rate in South Africa was reported 53% from 2016 to 2019 [18]. The procedures of handling samples from polio patients in the laboratory in South Africa followed the WHO Polio Laboratory Manual and supplement [18] and its sensitivity of polio detection was reported to be 0.97 [19].

We assumed monthly sampling (every 30 days) from the ES site in each patch for our simulation. Most African countries use a grab method to collect samples from wastewater and the sampling frequency is often 30 days due to manual labour burdens [20]. Some countries adopted a composite method for the ES, allowing daily or weekly sampling. GPEI recommends daily or weekly sampling but also accepts monthly sampling [21].

**Table S5.** Model Parameters used for our simulation.

| Parameter description | Values | Reference |
| --- | --- | --- |
| Per-dose vaccine effectiveness for IPV assessed by seroconversion | 0.63 | [22–24] |
| Average effective immune proportion (EIP) weighted by population size of children under 5 years old | 91.9% | Estimated from the population data (WorldPop) and vaccine coverage data |
| Basic reproduction number, $R_0$ | 14 | Assumed |
| Effective reproduction number at time 0 at a national level, $R_e(0)$ | 1.13 | Estimated |
| Latent period, $\gamma_1$ | 1/4 days<sup>-1</sup> | [25] |
| Infectious period, $\gamma_2$ | 1/15.02 days<sup>-1</sup> | Estimated [12] |
| Transmission rate, $\beta$ | 0.93 | Estimated by $R_0 \gamma_2$ |
| Travelling rates, $\alpha$ | 0.05 | Assumed |
| Age criteria for population | Less than 5 years old | Assumed |
| Birth and removal rate, $\mu$ | 1/(365*5) | Assumed |
| Transition rate of the incubation period for 6 compartments, $\sigma$ | 1/3.04 days<sup>-1</sup> | Estimated [5,13,14] |
| Patch-level ES population coverage, $p_c$ | 0.25 | Assumed |
| Paralysis-to-infection ratio, $p_{AFP}$ | 1/200 | [26] |
| Probability of seeking healthcare, $P_H$ | 0.9 | Assumed |
| Probability of stool sampling when a patient visits a hospital, $P_{AFP, sample}$ | 0.53 | [18,27] |
| Sensitivity of poliovirus detection when stool is tested, $P_{AFP, test}$ | 0.97 | [19,28] |
| Sensitivity of poliovirus testing of samples from sewage water, $P_{ES, test}$ | 0.97 | [19,28] |
| ES sensitivity | LogNormal($\mu=0.82, \sigma=1.45$) | Estimated [16] |
| Sampling frequency | Every 30 days | [20,21] |

### 1.4 Outcome measures

The detection pattern for each simulation falls into one of the following five patterns (Fig 1).

1. No detection: neither AFP surveillance nor ES detected the poliovirus circulation.
2. AFP surveillance only detected polio patients.
3. AFP surveillance detected the poliovirus circulation earlier than ES.
4. ES detected the poliovirus circulation earlier than AFP surveillance.
5. ES only detected the poliovirus circulation.

Let *t*_AFP_ and *t*_ES_ be the timing of the first detection through AFP surveillance and ES, respectively. The lead time of the first detection through ES over APF surveillance (denoted as *LT*) is defined as *t*_AFP_ – *t*_ES_, meaning that a positive value of *LT* corresponds to the early detection by ES (pattern 4) and a negative value of *LT* corresponds to the early detection by AFP surveillance (pattern 3). The lead time can only be calculated for patterns 3 and 4.

We used the proportion of each detection pattern among simulations with any poliovirus detection as the main outcome and excluded no detection pattern to match simulation outcomes with the real-world observations. To inform the quantitative aspect of early detection, we further classified pattern 3 into “< −60 LT” and “-60 ∼ −1 LT” categories and pattern 4 into “0 – 59 LT”, and “≥60 LT” categories. We refer to the “simulated early detection probability” as the proportion of “0 – 59 LT”, “≥60 LT” and ES only detection patterns given the poliovirus circulation is detected.

### 1.5 Average minimum distance to ES-covered patches and simulated early detection probability

We quantified the simulated early detection probability varying the number and location of the ES. However, our stochastic simulation took a long time and implementation was complex. We therefore explored a simple alternative measurement to inform the ES site layout strategy. Given importation risk distribution and EIPs in each patch were known, we calculated the weighted average minimum distance to the closest ES-covered patch and quantified the relationship between this measure and the simulated early detection probability.

The average minimum distance to the closest ES-covered patches (*d*_ave_) weighted by the importation risks is given as

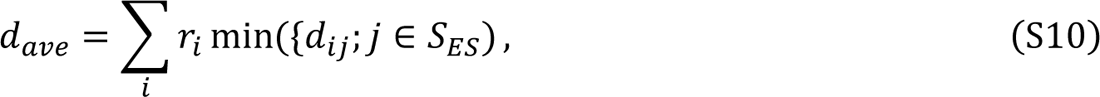

where *r*_i_ is the importation risk at patch *i* for each scenario, *d*_ij_ is the distance between patch *i* and patch *j*, and *S*_ES_ is the set of patches with the ES.

Since the regional heterogeneity in vaccination coverage can influence the timing of detection, we also consider the average minimum distance weighted by the importation risks and the outbreak probability in each patch (*d*_ave,w_):

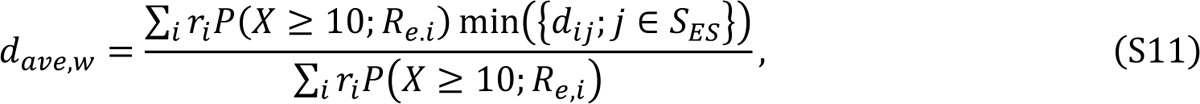

where *P*(*X* ≥ 10; *R*_*e,i*_) denotes the outbreak probability of 10 or more infections occurring given the effective reproduction number at patch *i* at the beginning of the simulation (*R*_e,i_), which is given by the product of the effective immunisation proportion at patch *i* (EIP_i_) and the basic reproduction number (*R*_0_). We visualised the relationship between *d*_ave,w_ and the simulated early detection ability in the main analysis (Fig 5). We arbitrarily chose the value of 10 infections for the cutoff. Assuming the branching process and the Poisson distribution as the offspring distribution (i.e. no overdispersion), the probability of observing x cases given *R*_e,i_ follows the Borel-Tanner distribution [29–31].

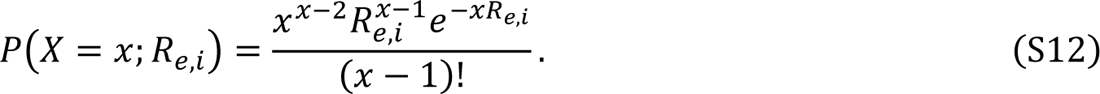

Then, the probability of 10 or more infections occurring is calculated by considering the complement of less than 10 infections occurring.

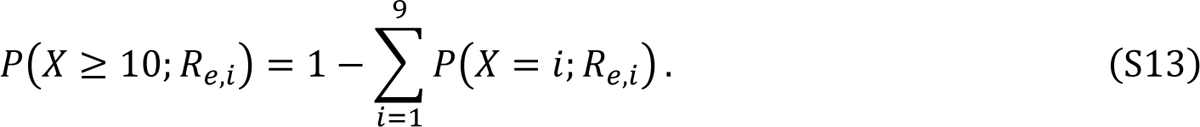

The histogram of *R*_e,i_ in South Africa and the corresponding probability of at least 10 infections happening given a single introduction is shown in Fig S9.

**Fig S9.**
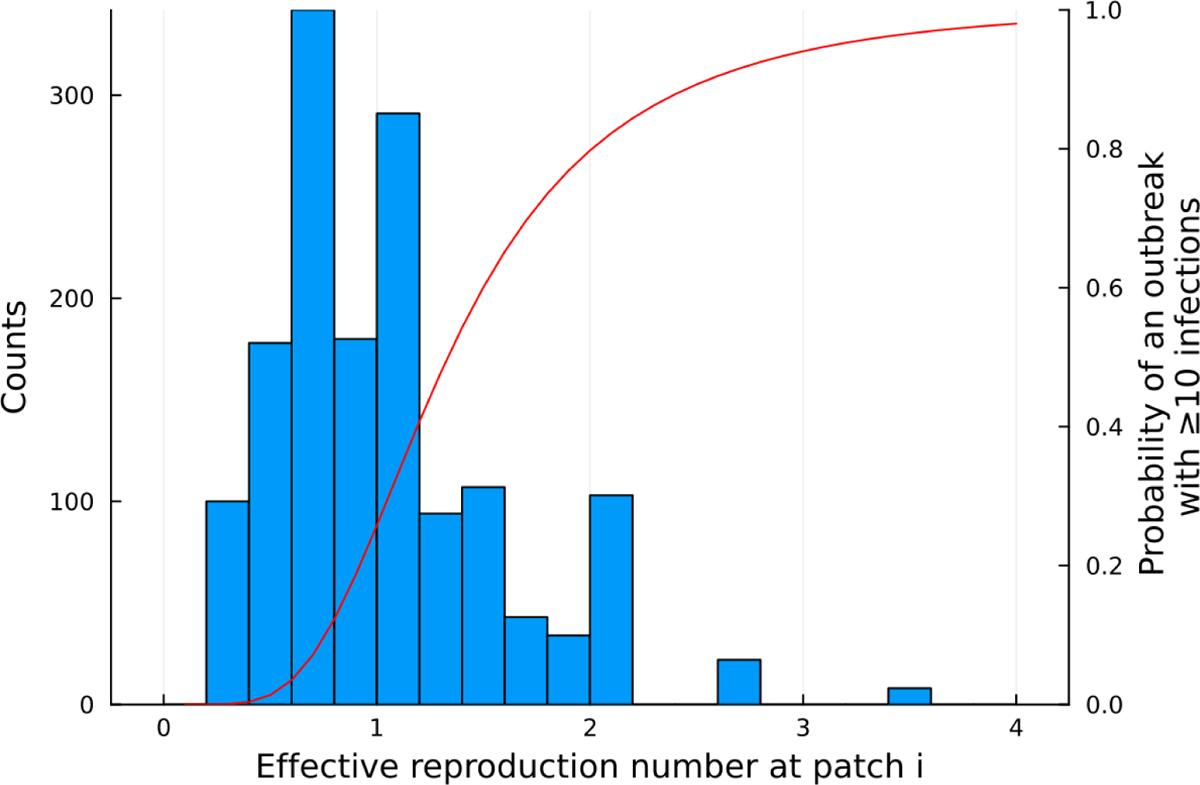
Histogram of the effective reproduction number for each patch and corresponding outbreak probability with ≥10 infections.

### 1.6 Simulation implementation

We randomly chose a patch for an introduction of WPV1 and simulated the stochastic meta-population model for 3 years 10,000 times for each scenario. We recorded the number of individuals at each patch in every compartment every day. Using those numbers in each compartment, we repeatedly applied the detection process through AFP surveillance and ES, varying the number of ES sites. We ended the simulation when one of the following criteria was met: the first poliovirus was detected through both surveillance, no polio patients were present, or three years had passed from the beginning of the simulation.

All the analysis was performed in Julia v1.8.3. It took around 5 to 8 hours to complete 10,000 simulations for one scenario from running the transmission model to running AFP surveillance model and ES model. The data and code are deposited on GitHub (https://github.com/toshiakiasakura/polio_environmental_surveillance).

## 2 Supplementary results

### 2.1 Characteristics for the top 20 populous patches

**Table S6.** Population size and effective immunisation proportion (EIP) for the top 20 populous patches.

| Longitude | Latitude | District of each patch | Population size aged under 5 years old | Cumulative proportion (%) | EIP<sup>d</sup> | Unimmunised population aged under 5 years old |
| --- | --- | --- | --- | --- | --- | --- |
| 27.77 | -26.15 | West Rand | 88395 | 3.1 | 94.9 | 4508 |
| 18.57 | -33.82 | City of Cape Town | 66488 | 5.5 | 94.7 | 3516 |
| 30.83 | -29.79 | eThekweni<sup>a</sup> | 55556 | 7.4 | 90.3 | 5408 |
| 28.15 | -25.96 | City of Johannesburg | 51209 | 9.2 | 93.4 | 3369 |
| 27.96 | -26.15 | City of Johannesburg | 48254 | 10.9 | 93.4 | 3175 |
| 30.83 | -29.60 | iLembe | 46696 | 12.6 | 75.4 | 11483 |
| 18.57 | -34.01 | City of Cape Town<sup>b</sup> | 40666 | 14.0 | 94.7 | 2151 |
| 18.37 | -33.82 | City of Cape Town | 40057 | 15.4 | 94.7 | 2118 |
| 27.96 | -25.96 | City of Johannesburg | 37234 | 16.7 | 93.4 | 2450 |
| 27.96 | -25.39 | City of Tshwane | 35821 | 18.0 | 94.5 | 1983 |
| 28.15 | -26.15 | Ekurhuleni<sup>c</sup> | 35621 | 19.3 | 94.1 | 2110 |
| 25.47 | -33.82 | Nelson Mandela Bay | 34878 | 20.5 | 91.5 | 2962 |
| 27.77 | -26.34 | West Rand | 27991 | 21.5 | 94.9 | 1427 |
| 27.96 | -25.77 | City of Tshwane | 25726 | 22.4 | 94.5 | 1424 |
| 28.15 | -25.58 | City of Tshwane | 25023 | 23.3 | 94.5 | 1385 |
| 27.77 | -26.54 | Sedibeng | 24300 | 24.1 | 91.9 | 1965 |
| 27.77 | -25.96 | West Rand | 20628 | 24.9 | 94.9 | 1052 |
| 28.34 | -25.58 | City of Tshwane | 19920 | 25.6 | 94.5 | 1103 |
| 28.34 | -26.15 | Ekurhuleni | 19597 | 26.3 | 94.1 | 1161 |
| 18.37 | -34.01 | City of Cape Town | 17993 | 26.9 | 94.7 | 952 |

<sup>a</sup> Closest patch to King Shaka International Airport among the patches in this table. The rank of population size in King Shaka International Airport's patch is 61<sup>st</sup>.
<sup>b</sup> Cape Town International Airport is in this patch.
<sup>c</sup> Tambo International is in this patch.
<sup>d</sup> Effective immunisation proportion.

### 2.2 Simulation results under a single patch setting

We simulated a stochastic SEIR model in a single patch to differentiate between the effects of parameters on the simulated early detection ability of ES attributable to spatial components and those stemming from model behaviours in a single patch. We employed the same model without a meta-population framework using the parameters listed in Table S5. We chose 100,000 population size of children aged under 5 years old and set the population size of all ages as 100,000 divided by the patch-level ES population coverage (*p*_c_).

We visualised the simulated cumulative detection probability over time for the AFP surveillance (dotted lines) and ES (solid lines) in Fig S10. Additionally, we visualised the proportion of any detection of the poliovirus circulation among all simulations including no detection pattern, and the proportion of each detection pattern given poliovirus circulation is detected (excluding no detection pattern).

Theoretically, ES-related parameters (sampling frequency, ES sensitivity, patch-level ES population coverage) do not influence the simulated cumulative probability of detection for AFP surveillance, which was confirmed by our simulations. Regarding population size, as the population size of children under 5 years old increased, the simulated cumulative probability for AFP surveillance also increased since the absolute value of patients with poliovirus became larger. On the other hand, the simulated cumulative probability for ES decreased because we assumed ES sensitivity was dependent on the incidence rate and a larger denominator resulted in the lower incidence rate.

We re-estimated ES sensitivity parameters with the simulated first detection timing through ES using Equation S5 to validate our assumption that ES sensitivity is proportional to the incidence rate of infectious individuals (Fig S11). We did not include simulated negative environmental samples for the estimation. Through this procedure, we illustrated higher ES sensitivity estimates for a large population size and lower ES sensitivity estimates for a small population size. This model behaviour is consistent with other estimates using the wastewater sample data [32].

**Fig S10.**
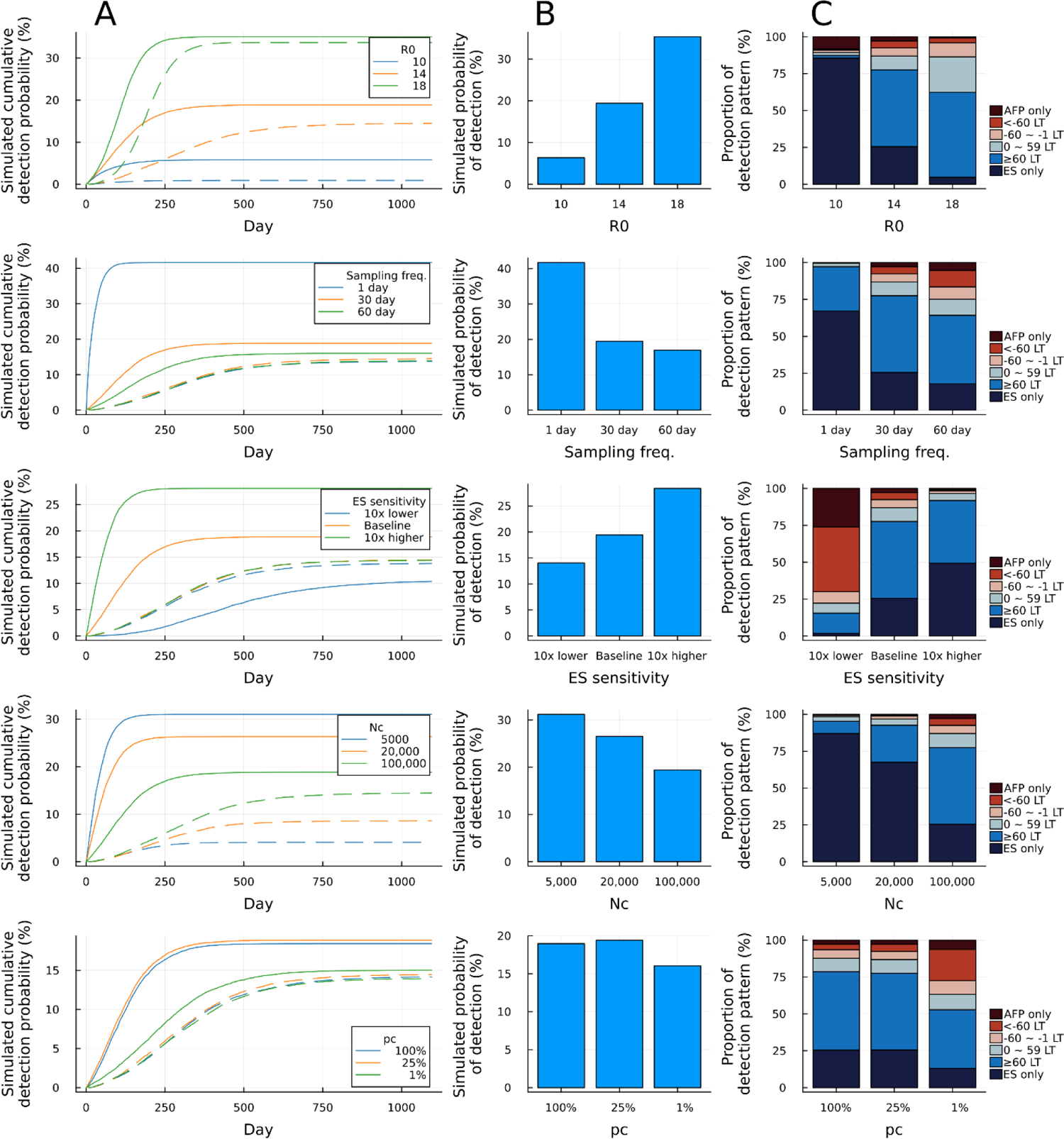
Sensitivity analysis of parameters in a single patch setting. (A) Simulated cumulative detection probabilities. Solid lines and dotted lines represent simulated probability for ES and AFP surveillance, respectively. (B) Simulated probability of detection through either AFP surveillance or ES among all simulations including no detection pattern (%). (C) The proportion of each detection pattern (%) given poliovirus circulation was detected.

**Fig S11.**
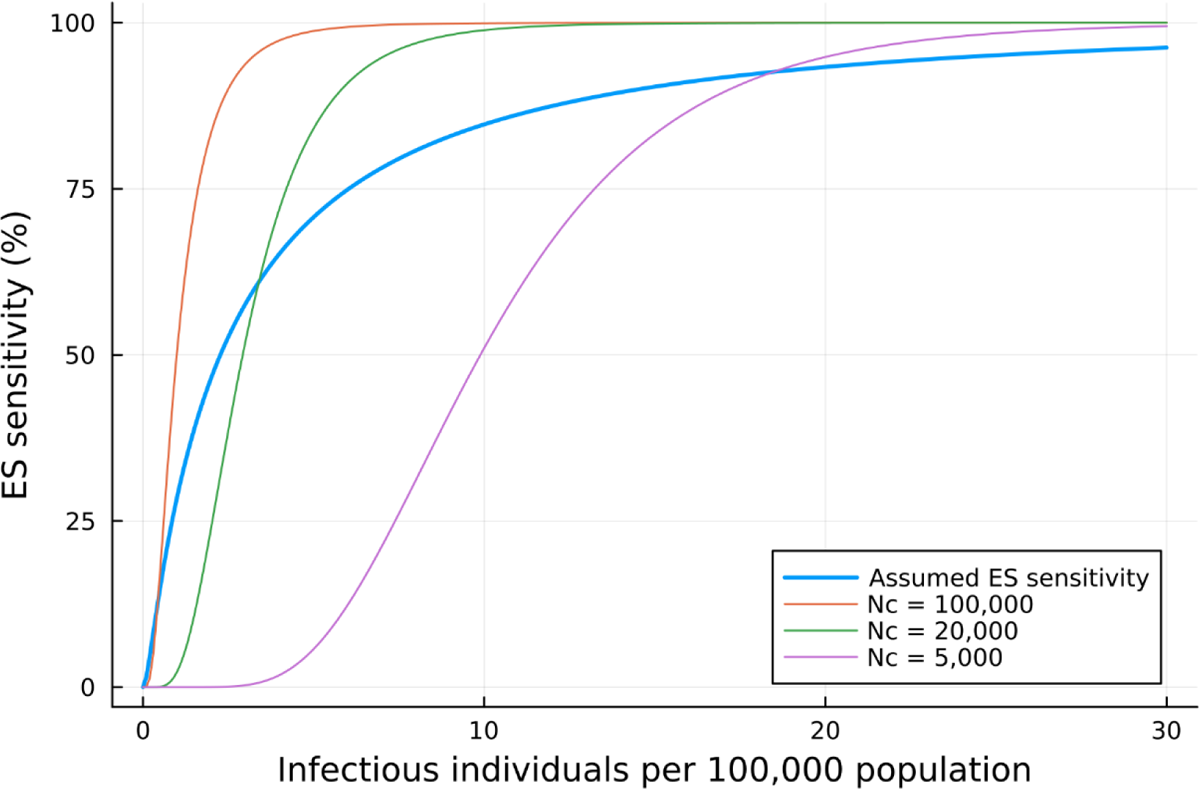
Dose-response curves for estimated ES sensitivity parameters under different population sizes of children under 5 years old (*N*_c_) in a single patch setting.

### 2.3 Two alternative visualisations for the main analysis

We limited the x-axis for Fig 2 to 160 for the interpretability of the results, and here we visualised the same results where the x-axis represents the national ES population coverage (Fig S12). The national ES population coverage was obtained by the product of the percentage of the population in ES-covered patches and the patch-level ES population coverage (*p*_c_). Therefore, the maximum value of the ES population coverage is matched with our parameter choice for *p*_c_ (i.e. 25%). It is noted that the maximum number of ES-covered patches of 1502 corresponds to the national ES population coverage of 25%, and 160 ES-covered patches were close to the plateau of the simulated early detection probability.

We further calculated the proportion of each detection pattern by including the “no detection” pattern (Fig S13) whereas we did not include the “no detection” pattern in the main text figures. Differences in the proportion of the “no detection” pattern would be attributable to the ES layout strategy and heterogeneity in the effective reproduction number in each path (Fig S9).

**Fig S12.**
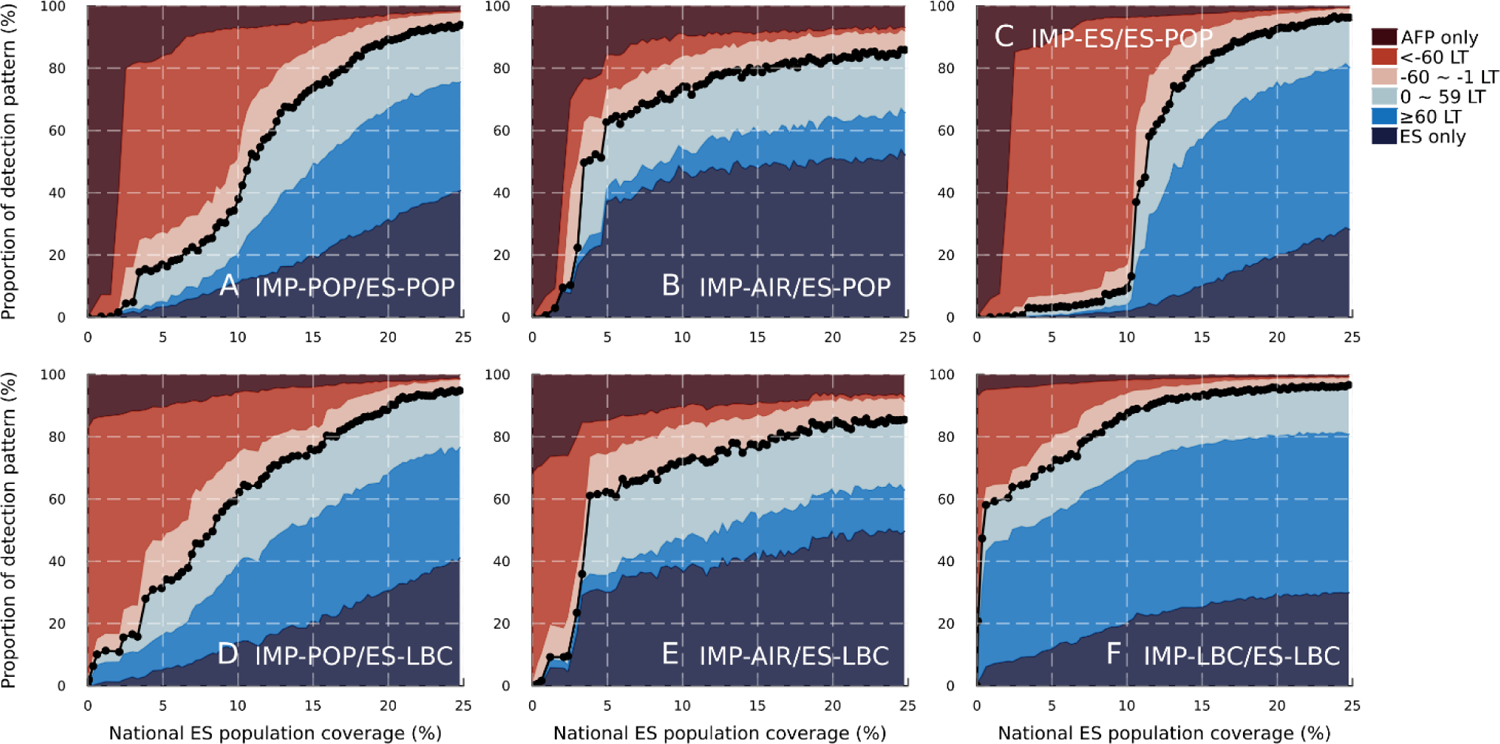
Proportion of each detection pattern (%) against national ES population coverage for 6 scenarios. The blue-coloured area under the black dotted lines represents the simulated early detection probability, consisting of early detection of ES over AFP surveillance and ES only detection. LT denotes the lead time of poliovirus detection through ES over AFP surveillance.

**Fig S13.**
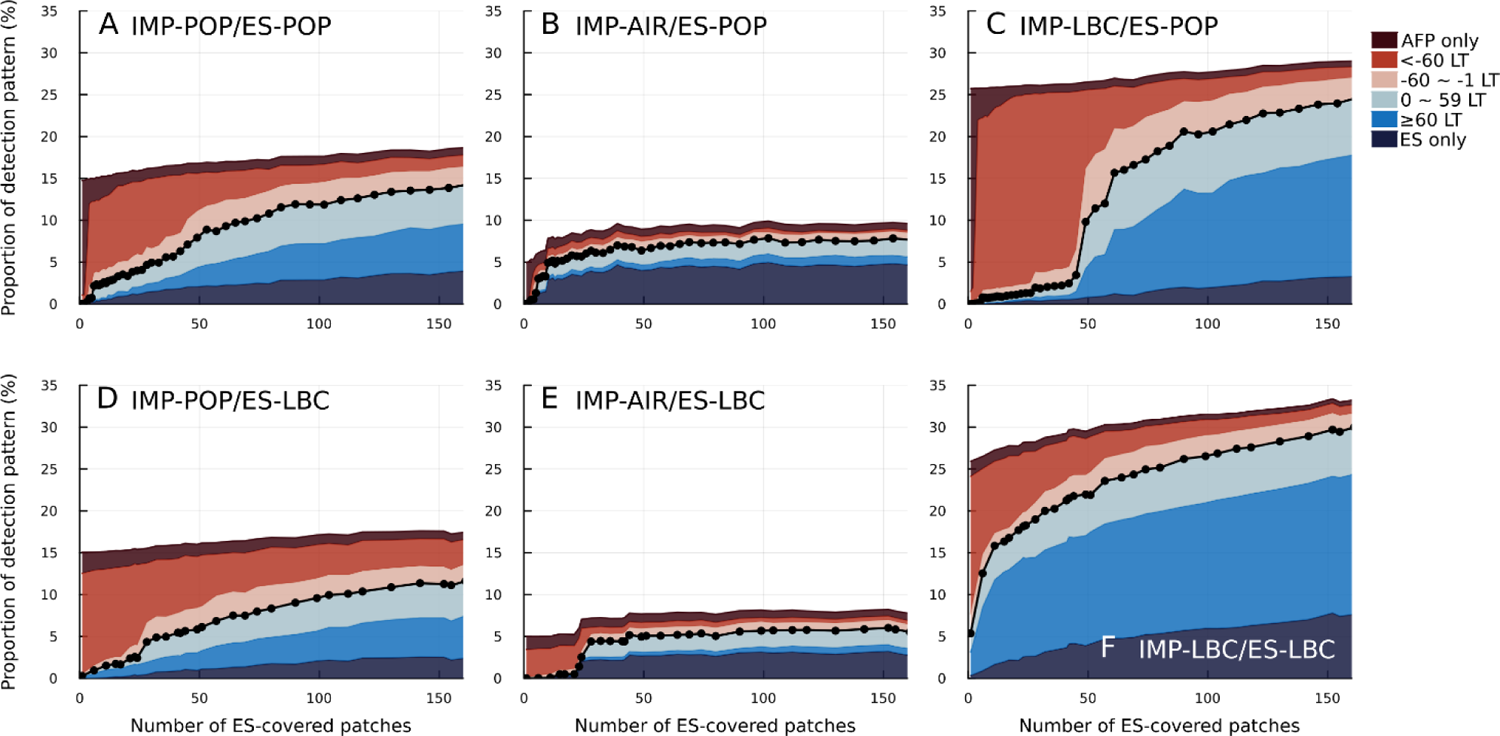
Proportion of each detection pattern (%) including the no detection pattern against the number of ES-covered patches for 6 scenarios. The non-coloured area represents the simulated probability of detecting poliovirus circulation neither through AFP surveillance nor ES. The blue-colours area under the black dotted lines represents the simulated early detection probability. LT denotes the lead time of poliovirus detection through ES over AFP surveillance.

### 2.4 Sensitivity analysis on the patch-level ES population coverage for different scenarios

We conducted the sensitivity analysis of the patch-level ES population coverage (*p_c_*) for the other 5 scenarios as well as the main text (Fig 4). We found a similar trend in Fig S14-S15 as in Fig 4. That is, the simulated early detection probability was robust against the number of ES-covered patches but had a large variation against the national ES population coverage.

**Fig S14.**
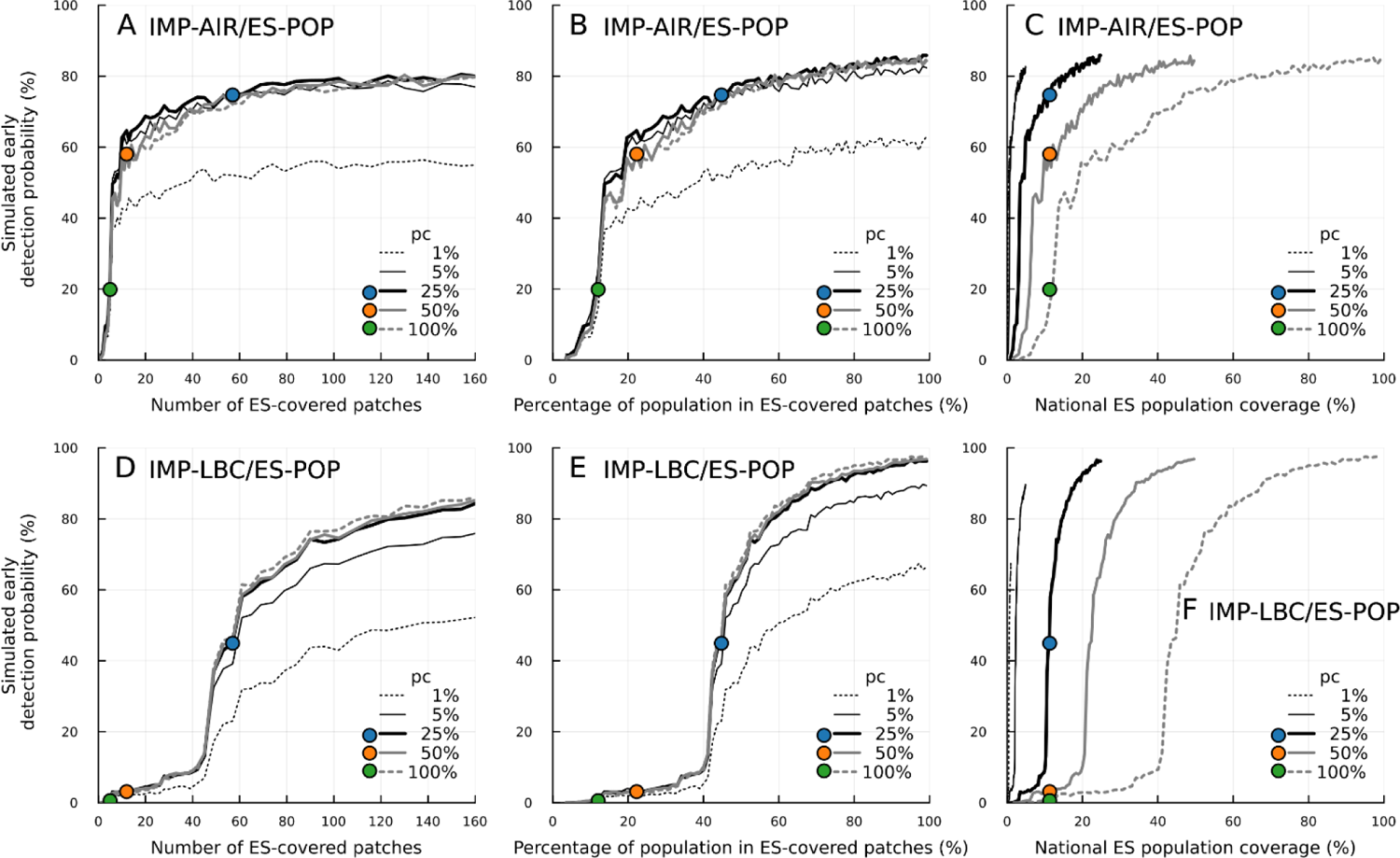
Sensitivity analysis of the patch-level ES population coverage, *p*_c_, for the ES-pop scenarios. (A, B, C) For the IMP-AIR/ES-POP scenario. (C, D, F) For the IMP-LBC/ES-POP scenario. Simulated early detection probability is plotted (A, D) against the number of ES-covered patches, (B, E) against the percentage of the population in ES-covered patches, and (C, F) against the national ES population coverage. The data points represent simulations where the national ES population coverage of the simulated ES layout aligns with the current coverage in South Africa (11.3%), under *p_c_* of 25% (blue), 50% (orange) and 100% (orange). The national ES population coverage is given by the product of *p*_c_ and the percentage of the population in ES-coverage patches. It is noted that the maximum number of ES-covered patches is 1502 and the x-axis for (A, D) is limited to a maximum value of 160.

**Fig S15.**
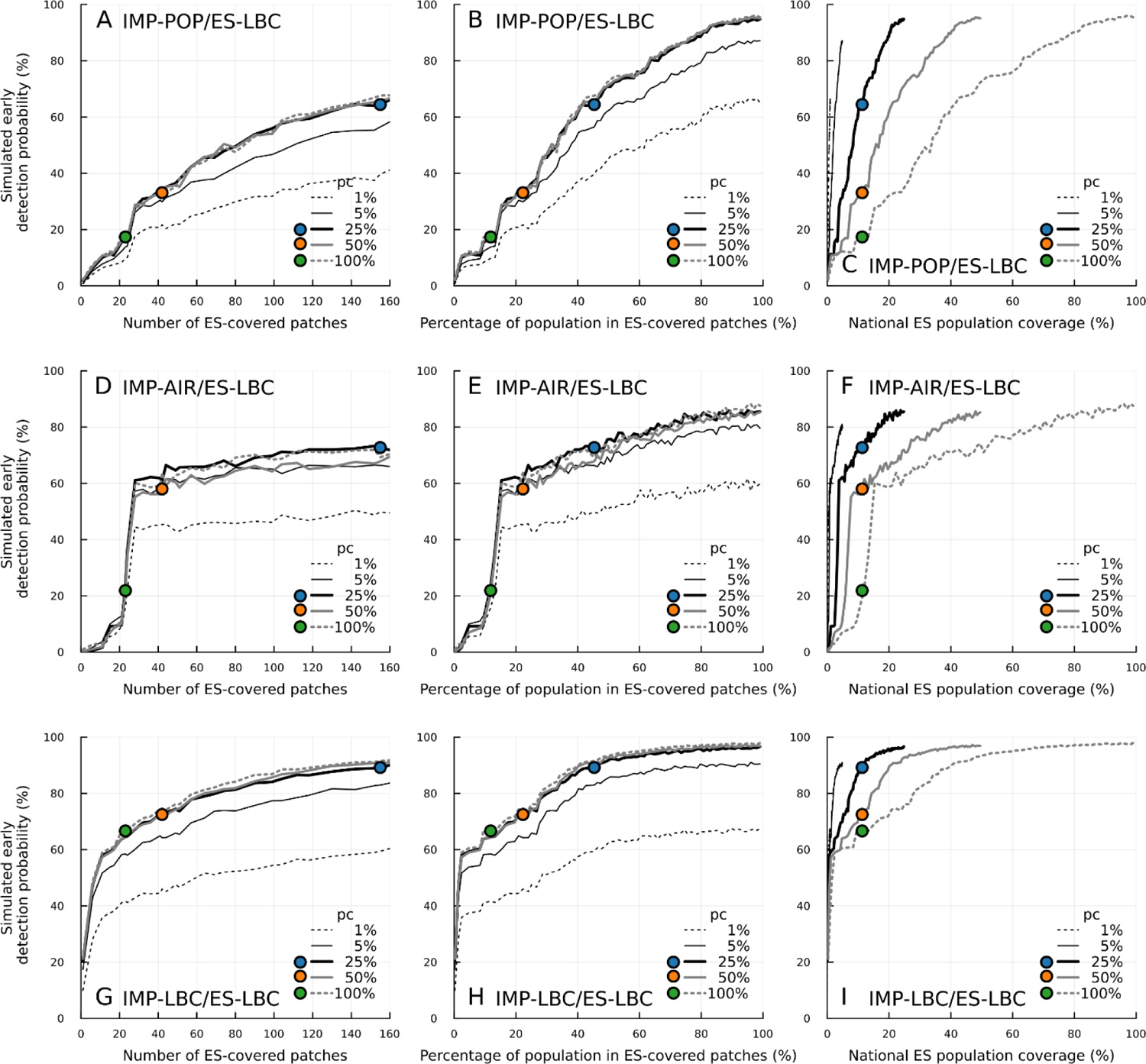
Sensitivity analysis of the patch-level ES population coverage, *p*_c_, for the ES-LBC scenarios. (A, B, C) For the IMP-POP/ES-LBC scenario. (D, E, F) For the IMP-AIR/ES-LBC scenario. (G, H, I) For the IMP-LBC/ES-LBC scenario. Simulated early detection probability is plotted (A, D, G) against the number of ES-covered patches, (B, E, H) against the percentage of the population in ES-covered patches, and (C, F, I) against the national ES population coverage. The data points represent simulations where the national ES population coverage of the simulated ES layout aligns with the current coverage in South Africa (11.3%), under *p_c_* of 25% (blue), 50% (orange) and 100% (orange). The national ES population coverage is given by the product of *p*_c_ and the percentage of the population in ES-coverage patches. It is noted that the maximum number of ES-covered patches is 1502 and the x-axis for (A, D, G) is limited to a maximum value of 160.

---

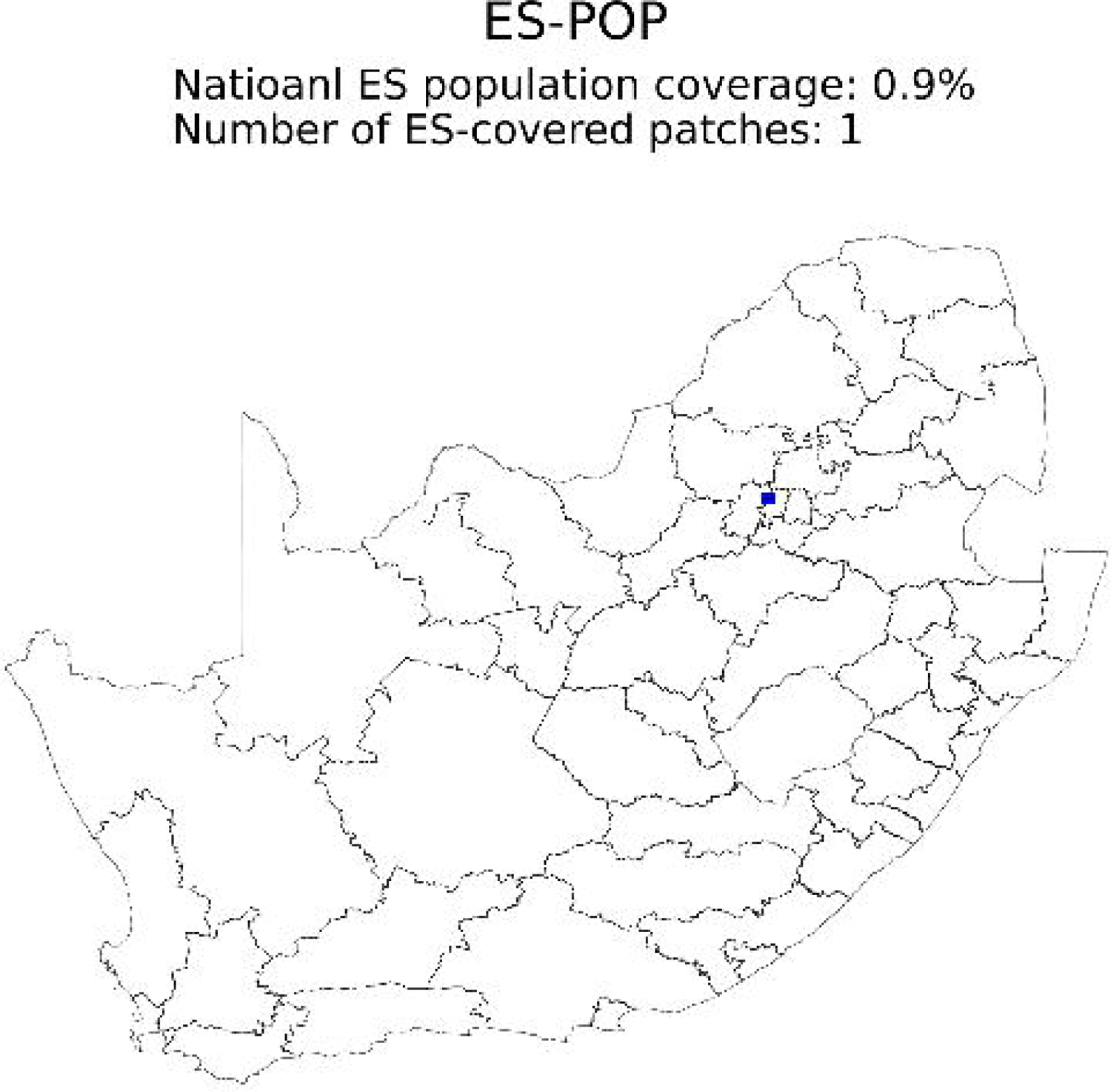

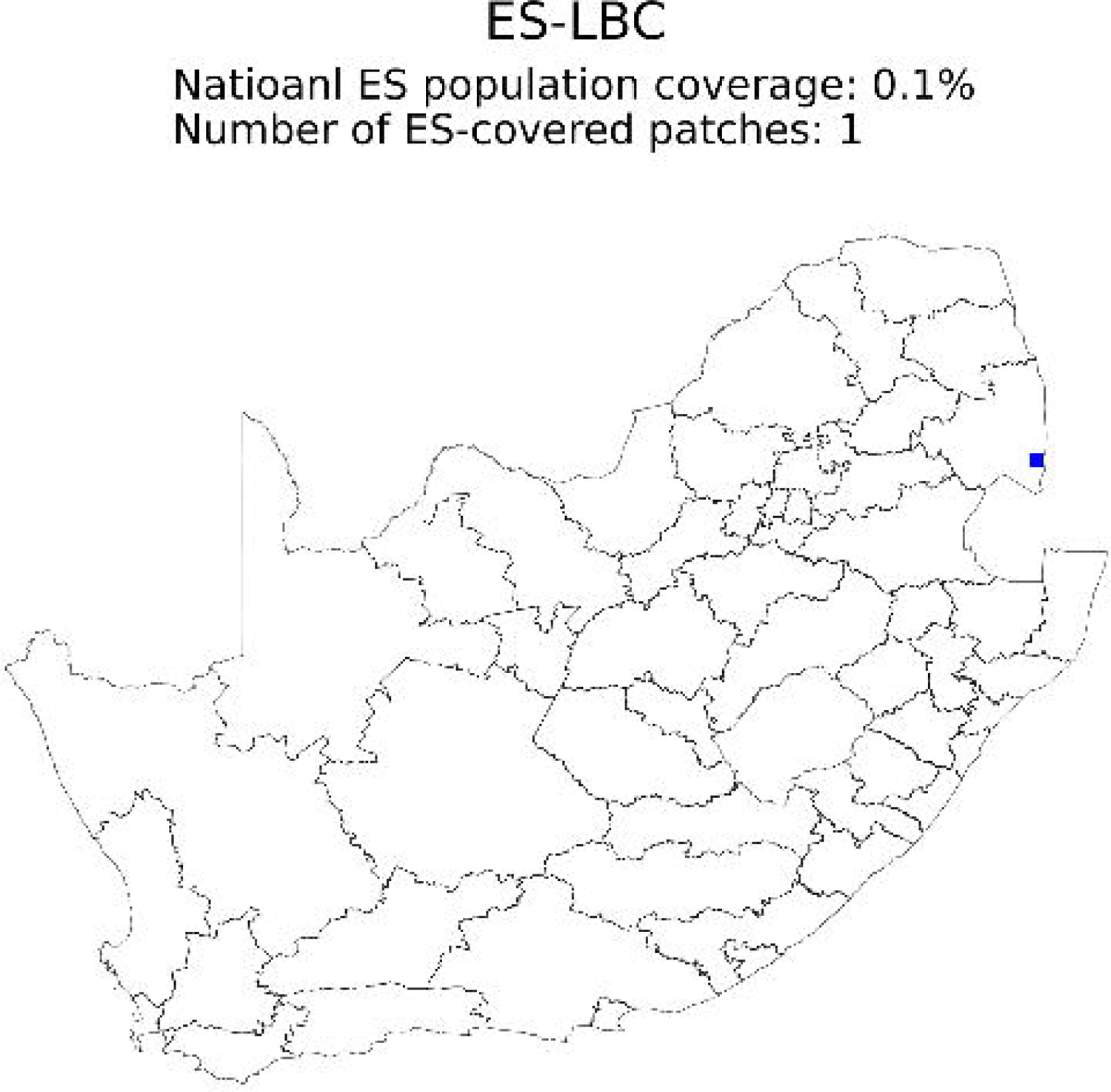

